# Computational Modeling of Ovarian Cancer Reveals Optimal Strategies for Therapy and Screening

**DOI:** 10.1101/19009712

**Authors:** Shengqing Gu, Stephanie Lheureux, Azin Sayad, Paulina Cybulska, Liat Ben-David Hogen, Iryna Vyarvelska, Dongsheng Tu, Wendy Parulekar, Matthew Nankivell, Sean Kehoe, Dennis Chi, Douglas A. Levine, Marcus Q. Bernardini, Barry Rosen, Amit Oza, Benjamin G. Neel

## Abstract

High-grade serous tubo-ovarian carcinoma (HGSC) is a major cause of cancer-related death. Whether treatment order—primary debulking surgery followed by adjuvant chemotherapy (PDS) or neo-adjuvant chemotherapy with interval surgery (NACT)—affects outcome is controversial. We developed a mathematical framework that holds for hierarchical or stochastic models of tumor initiation and reproduces HGSC clinical course. After estimating parameter values, we infer that most patients harbor chemo-resistant HGSC cells at diagnosis, and that if complete debulking (<1 mm residual tumor) can be achieved, PDS is superior to NACT due to better depletion of resistant cells. We further predict that earlier diagnosis of primary HGSC, followed by complete debulking, could improve survival, but its benefit in relapsed patients is likely to be limited. Our predictions are supported by primary clinical data from multiple cohorts. Our results have clear implications for these key issues in HGSC management.

**Significance Statement:** The optimal order and timing of surgery and chemotherapy, and the potential benefits of earlier diagnosis of HGSC, remain controversial. We developed a mathematical framework of tumor dynamics to address such issues, populated the model with primary clinical data and reliably recapitulated clinical observations. Our model prospectively predicts that: (1) PDS is superior to NACT when complete debulking is feasible; (2) timely adjuvant chemotherapy is critical for the outcome of PDS with <1mm, but not >1mm, residual tumors; (3) earlier detection of relapse is unlikely to be beneficial with current therapies; (4) earlier detection of primary HGSC, followed by complete debulking, could have substantial benefit. Our model provides insights into the evolutionary dynamics of HGSC, argues for new clinical trials to optimize HGSC therapy, and is potentially applicable to other tumor types.

## Introduction

Ovarian cancer is the 8^th^ most common cancer in women and the 8^th^ most common cause of cancer death in women worldwide^1^. High-grade serous ovarian cancer (HGSC) constitutes ~70% of all ovarian malignancies and has the worst prognosis^2^. Current treatment of HGSC consists of cytoreductive surgery and combination chemotherapy with platinum-containing DNA-crosslinking drugs and taxane-based microtubule-stabilizing agents^2^. Although treatment significantly improves survival, most women with HGSC relapse with chemotherapy-refractory disease and eventually succumb^3^. Multiple mechanisms of chemo-resistance have been documented^4,5^, including reduced intracellular drug accumulation^6^, detoxification by increased levels of glutathione^7^, altered DNA damage repair^8,9^, dysfunctional apoptotic pathways^10,11^, and hyper-activation of various cell signaling pathways^12-14^. These mechanistic studies are consistent with recent genomic analyses that reveal marked clonal evolution of HGSC during therapy^15^. Other evidence, however, supports a hierarchical organization of HGSC, featuring intrinsically chemo- resistant “cancer stem cells” (CSC) that can escape initial treatment and seed recurrence^16-18^.

Although there is uniform agreement that HGSC patients should receive surgery and chemotherapy, there is disagreement over the optimal order and timing of these modalities. Two main options exist: primary debulking surgery with adjuvant chemotherapy (PDS), or neo-adjuvant chemotherapy, followed by interval debulking surgery (NACT)^19-24^. In either case, the surgical standard of care is to seek maximal cytoreduction, with the objective being to leave no visible residual disease. However, the precise definition of such “optimal debulking” varies among different centers, surgeons, and reports^19,21,24,25^.

Several studies have reported similar outcomes after PDS or NACT, including two highly influential randomized trials (EORTC and CHORUS) carried out across multiple countries^22,23,26-28^. In both trials, however, there was a bias in patient recruitment, favoring those with more extensive disease, who are less likely benefit from “upfront” surgery^23,28^. Consistent with this interpretation, overall survival in these trials was significantly shorter than that seen in other HGSC cohorts^19,24,29,30^. Closer examination of these studies revealed additional factors that might have influenced their conclusions. The EORTC study had inconsistencies in optimal debulking rates between participating centers, with the PDS- associated complete debulking data highly influenced by the results from a single institute^23^. The CHORUS study involved 76 clinical sites, and there were substantial differences in surgery execution and chemotherapy drug selection/dosage between those institutions^28^.

A review of cases at our center (Princess Margaret Cancer Center) revealed that PDS patients with no visible disease post-resection survived substantially longer (7-year survival >60%) than those receiving NACT (7-year survival ~10%). Furthermore, although residual tumor post-resection is a critical determinant of survival, its influence on the PDS group was far more dramatic than on NACT group^24^. Of course, this report suffers from deficiencies common to all retrospective analyses, including lack of randomization to account for tumor burden at diagnosis and other factors; indeed, the NACT group in this study did have more extensive disease.

Another controversy in HGSC management focuses on the potential benefit of earlier diagnosis. Earlier diagnosis of primary HGSC is generally assumed to enhance patient survival and quality of life^3^. Intuitively, one might predict that the same reasoning would apply to recurrent disease; however, survival reportedly is similar in relapsed patients treated earlier, based on increasing serum CA125 levels, versus those treated only when physical symptoms of recurrence appear^31^. Conceivably, the lead time between CA125 rise and clinical recurrence is too short for earlier chemotherapy to be beneficial; if so, then patient survival might be extended by more sensitive methods, such as testing for circulating tumor DNA (ctDNA)^32,33^.

To address these issues, we developed a mathematical framework that models the dynamics of HGSC progression, response to surgery and chemotherapy, and recurrence. Our results, generated over a wide range of parameters and accounting for hierarchical and stochastic models of tumor initiation, argue that PDS is superior to NACT when complete debulking is feasible and suggest that with currently available therapies, the benefits of earlier detection are intrinsically restricted to primary HGSC.

## Results

### Clinical Cohorts

To train and evaluate our model, we used two independent cohorts, comprising a total of 285 FIGO Stage IIIC-IV HGSC patients (Table 1 and Figure S1). The first dataset contains information on 148 patients who were treated by PDS or NACT at University Health Network (UHN) between March 2003 and November 2011^24^. In this cohort, “complete (microscopic) debulking” (<1mm residual tumor) was achieved in 97 patients, whereas 51 patients had 1-10mm of residual tumor. Among the 97 patients with complete debulking, 40 received upfront surgery, followed by 6 cycles of platinum and taxane-based chemotherapy (hereafter, “PDS <1mm”), whereas the other 57 patients were treated with upfront chemotherapy, followed by interval surgery after the first 3-4 cycles (hereafter, “NACT <1mm”). For each patient, descriptions of tumor size post-surgery and serum CA125 levels over the course of treatment were recorded (Figure 1a). As reported previously^24^, overall survival was much better for patients in the PDS <1mm group (Figure 1b). However, patients in the NACT <1mm group tended to have greater tumor burden at diagnosis^24^, confounding direct comparison of the survival curves. Data from these patients (termed “Training Set” below) were used to train the parameters in our computational model.

**Table 1.** Patient Characteristics in UHN and NCIC Cohorts.

|  |  | UHN Cohort | NCIC Cohort |
| --- | --- | --- | --- |
| Total number of patients |  | 148 | 137 |
| Time of diagnosis* |  | 2008 JAN<br>(2005 OCT – 2009 MAR) | 2003 NOV<br>(2003 JAN – 2004 AUG) |
| Age* |  | 58 (50 – 66) | 58 (52 – 64) |
| Treatment regimen | PDS | 61 (41%) | 137 (100%) |
|  | NACT | 87 (59%) | 0 (0%) |
| FIGO Stage | IIIC | 124 (84%) | 102 (74%) |
|  | IV | 24 (16%) | 35 (26%) |
| Residual disease | <1mm | 97 (66%) | 20 (15%) |
|  | 1-10mm | 51 (34%) | 34 (25%) |
|  | >10mm | 0 (0%) | 83 (60%) |
| CA125 before treatment* |  | 686 (220 – 1680) | Unknown |
| Overall survival* |  | 41 (22 – not reached) | 40 (26 – not reached) |

\* values indicate median (range of first and third quartiles).

**Figure 1.**
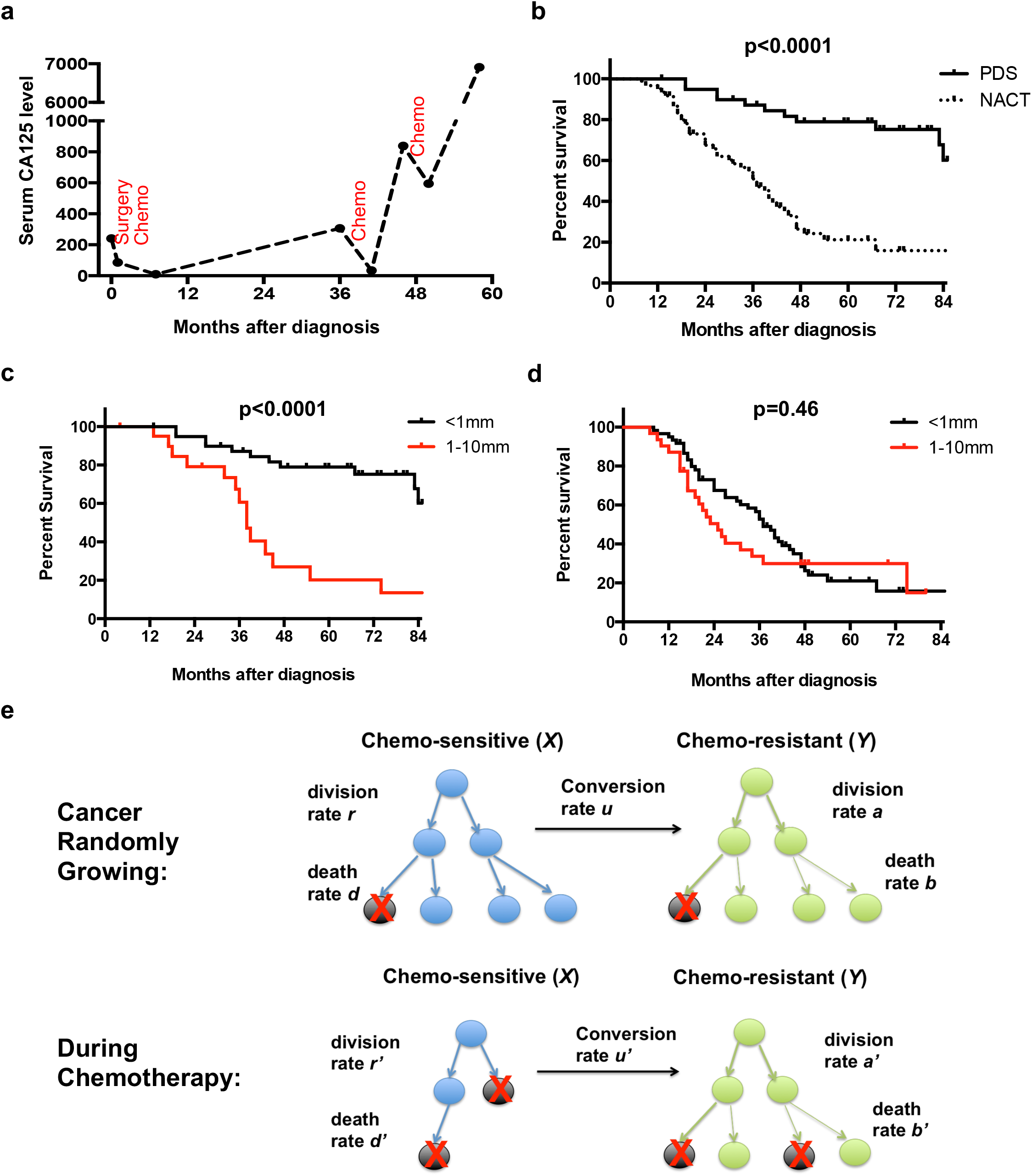
Mathematical Framework of HGSC Clinical Course. (a) Representative CA125 levels during clinical course of a typical HGSC patient. Chemotherapy responsiveness decreases along the treatment course, indicating the accumulation of chemo-resistant cells. (b-d) Kaplan-Meier survival curves for patients in the UHN dataset. Panel (*b*) compares the survival of patients with <1mm residual tumor after treatment by PDS or NACT. Note that patients with PDS <1mm lived significantly longer than those with NACT <1mm (p<0.0001). Panel (c) compares the survival of patients treated by PDS with <1mm versus 1-10mm residual tumor. Note that patients with PDS <1mm lived significantly longer than those with PDS 1-10mm (p<0.0001). Panel (d) compares the survival of patients treated by NACT with <1mm versus 1-10mm residual tumor. Note that survival of these two groups was not significantly different (p=0.46). (e) Mathematical framework for modeling HGSC progression. The model assumes the existence of chemo-sensitive and chemo-resistant HGSC cells. During random growth, chemo-sensitive cells divide at rate *r*, die at rate *d*, and convert into chemo-resistant cells with probability *u* per cell division; chemo-resistant cells divide at rate *a* and die at rate *b*. During chemotherapy, chemo-sensitive cells divide at rate *r*’, die at rate *d’*, and convert into chemo-resistant cells with probability *u*’ per cell division; chemo-resistant cells divide at rate *a’* and die at rate *b*’. See text and Supplementary Experimental Procedures for details.

The other 51 patients in the UHN dataset underwent debulking, but were left with residual tumor of 1-10mm in diameter. Incomplete debulking usually was due to tumor location. As reported previously^24^, there was a significant negative association between residual tumor size and patient survival in patients who had undergone PDS (Figure 1c), but no association was found in the NACT group (Figure 1d). Similar trends have been observed in several other studies investigating the association between patient survival and residual tumor size^19,20,34^. Independent of the <1mm cohort, we used this 1-10mm cohort (“Validation Set 1”, below) in initial tests of the validity of our computational model (Figure S1).

The second dataset, which is independent of the UHN cohort, contains information on 137 patients diagnosed from October 2001 to June 2005 and enrolled in the CAN-NCIC- OV16 trial (hereafter, “NCIC cohort”). These patients all received upfront surgery, followed by 8 cycles of carboplatin/paclitaxel chemotherapy. The median delay for chemotherapy after surgery was 0.8 month (compared with 1 month at UHN). In 20 patients, debulking to <1mm residual tumor was achieved, 36 patients had 1-10mm residual tumor, and 81 patients had residual tumor diameter >10mm. Data from the NCIC cohort was used to further evaluate the validity of our computational model (Figure S1).

### Model Overview

We developed a mathematical model of cancer initiation and evolution to investigate the dynamics of HGSC growth, the onset of chemo-resistance, and the effects of various treatment strategies on patient survival (Figure 1e). Our initial framework was based on work by the Michor laboratory^35,36^, and considers exponential expansion of HGSC cells starting from a single cancer cell that has all of the genetic alterations needed for proliferation and metastasis, but has not developed chemo-resistance. This framework assumes that during tumor development/progression, any chemo-sensitive HGSC cell can acquire mutations and/or epigenetic alterations that enable chemo-resistance; i.e., we assume a stochastic model of tumor initiation. However, the model can be modified to accommodate a hierarchical organization of HGSC, in which a relatively chemo-resistant CSC gives rise to chemo-sensitive progeny. Importantly, the predictions/implications of the hierarchical and stochastic models are essentially the same (see below and Supplemental Information).

In the absence of chemotherapy, chemo-sensitive cells divide at rate *r* and die at rate *d*, genetic or epigenetic changes that cause chemo-resistance occur with probability *u* per cell division, and chemo-resistant cells divide at rate *a* and die at rate *b* (which can be the same or different as the rates of division and death of chemo-sensitive HGSC cells). The total number of HGSC cells at diagnosis is denoted by *M_1_*, with a proportion *εM_1_* metastasized outside the peritoneal cavity and not accessible at debulking surgery. Once cancer has been diagnosed, the (virtual) patient receives PDS or NACT. Post-surgery, the number of HGSC cells in the peritoneal cavity is reduced to *M*; cancer cells at inaccessible locations persist. During chemotherapy, chemo-sensitive cells divide at rate *r’* and die at rate *d’*, with *r’* < *d’*; genetic or epigenetic changes that cause chemo-resistance occur with probability *u*’ per cell division; and chemo-resistant cells divide at rate *a’* and die at rate *b*’. After first line treatment, persisting sensitive and resistant cells continue to proliferate and reach number *M_1_*, at which time relapse is diagnosed, and the patient is again treated with chemotherapy. Note that although tumor recurrence is usually diagnosed before it reaches *M_1_* in the clinic, our analyses (see below and Figure 6), along with clinical evidence^31^, suggest that the timing of diagnosis of recurrence does not affect treatment outcome. This regression-relapse cycle is repeated until chemotherapy fails to reduce disease burden, at which point tumor growth continues until the patient’s death, with a total number of cancer cells denoted by *M_2_*, which is larger than *M_1_*.

This framework enables us to estimate the probability that chemo-resistance is present at diagnosis, the dynamics of sensitive and resistant cell numbers along the treatment course, and the length of survival of HGSC patients. Analytic approximations for these quantities are shown in the Methods section. With these quantities, we estimated the conversion rate (*u*) and inaccessible proportion (ε) by minimizing the deviations between patient survival data and the corresponding predictions obtained using our model. Using these estimates, we then predicted the survival of patients in test datasets to assess the validity of our model, compared the survival of PDS- or NACT- treated patients when controlled for starting tumor burden, and probed the potential benefits of earlier diagnosis of HGSC.

### Estimation of Parameter Values

For most model parameters, a clinically relevant range of values could be deduced from clinical data or previous publications (see Methods section for detailed description of parameter value estimations). We then varied *u* and *ε* over a wide range, and computed the expected distribution of survival of patients by Monte-Carlo simulation. We compared the deviation between the Training Set data and the predictions of our model for each combination of *u* and *ε*, and from the region of best fit for these parameters, we inferred that 10^-9^ < *u* < 10^-7^ and 10^-9^ < *ε* < 10^-5^ (Figure 2a). Finer investigation of the fit between data and theory in this parameter region showed that *u* = 10^-76^ and *ε* = 10^-74^ is the best combination of these parameters, as it minimizes the deviation between data and theory (Figure 2b). Importantly, the trained value for *u* is consistent with chemo-resistant conversion rates estimated for multiple other cancers^37-40^. The trained value for *ε* predicts that inaccessible cancer cells outside the peritoneum are less numerous than cancer cells inside the peritoneum after first-line therapy, which comports with clinical observations that recurrent HGSC occurs predominantly inside the abdominal cavity. Using the trained parameter values, we compared the observed (Figure 1b) and predicted (Figure 2c) distributions of patient survival in the Training Set. Reassuringly, the model predictions closely recapitulated the clinical observations (Figures 2d-e). Notably, although this combination of *u* and *ε* represented the best prediction in the training set, and big deviations from these values led to worse predictions (Figure S2a-b), adjusting their values over a smaller range did not lead to substantial decrease in prediction accuracy (Figure S2c). Therefore, in our follow-up analyses, in addition to using the base values of *u* = 10^-76^ and *ε* = 10^-74^, we also tried alternative combinations to test the robustness of our model predictions.

**Figure 2.**
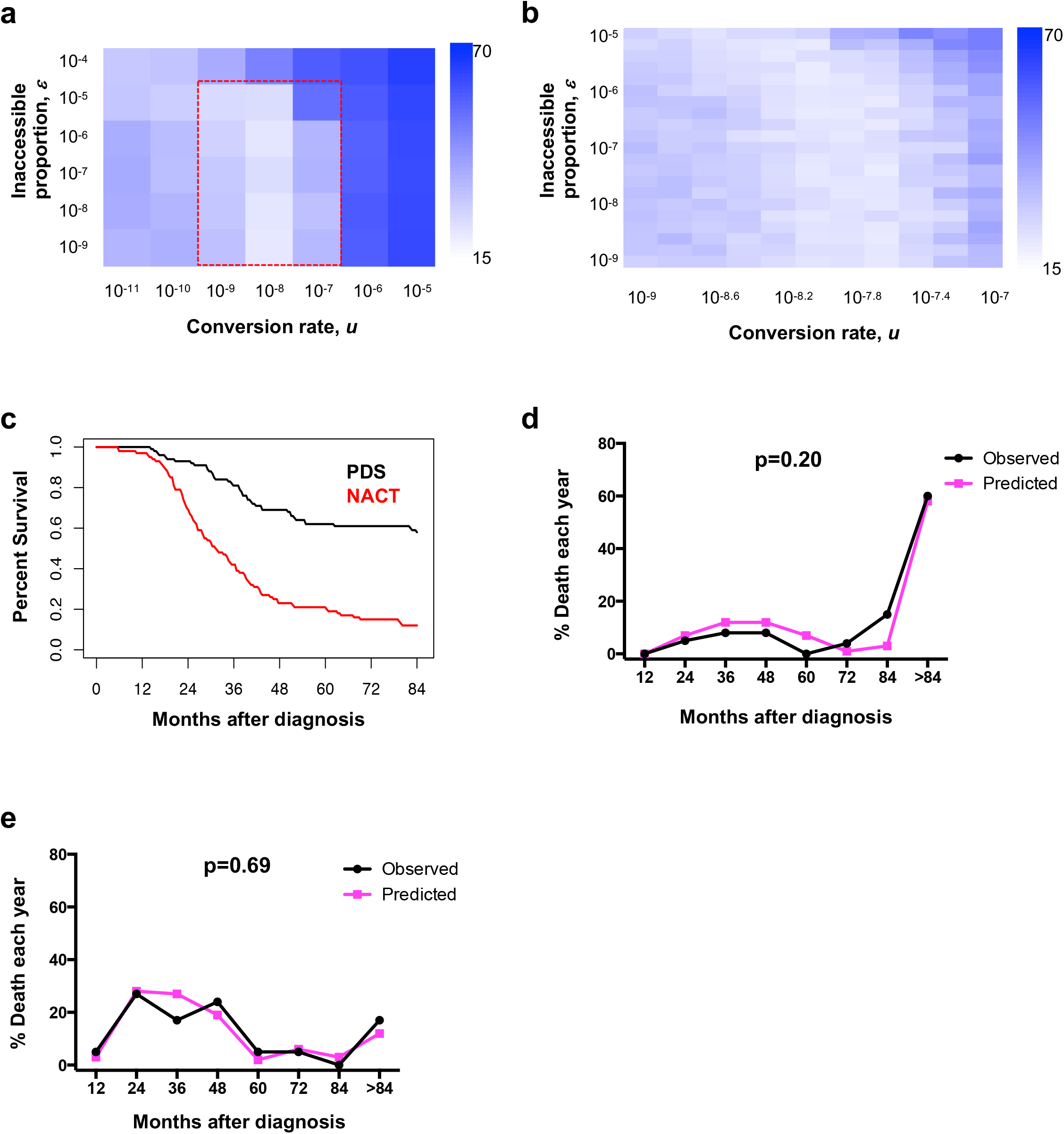
Estimation of Parameter Values and Evaluation of Model Predictions. (a-b) Estimation of conversion rate (*u*) and inaccessible proportion (*ε*) in patients treated by PDS or NACT with <1mm residual tumor, using data from the UHN cohort (Training Set). Colors represent the degree of deviation between clinical data and the predictions of the mathematical model. Lighter colors represent region of best fit between theory and observation. Panel (*b*) provides a finer scale analysis of the red dashed region in panel (a). (c) Model prediction of survival of patients treated by PDS or NACT with <1mm residual tumor. Parameter values are: *u* = 10^-7.6^, ε= 10^-7.4^, *d* = *r*/10, *b* = *a*/10, *a’* = *a*, *b*’ = *a*/5, *u*’ = 10*u*. Values for *r*, *a, r’*, and *d’* were obtained from normal distributions. *r* was set with mean 2 and s.d. 0.8. *a* was set with mean 0.84 and s.d. 0.42. *r’* was set with mean 0.2 and s.d. 0.08. *d’* was set with mean 4.9 and s.d. 1. *M_1_, M_2_*, and *M* were obtained from normal distributions in the log-10 scale. *log_10_M_1_* was set with mean 11.5 and s.d. 0.4 for PDS group, and mean 12 and s.d. 0.4 for NACT group. *log_10_M_2_* was set with mean 13 and s.d. 0.4. *log_10_M* was set with mean 6 and s.d. 0.4 for <1mm residual cancer. (d-e) Comparisons of predicted and observed overall survival for PDS <1mm (d) and NACT <1mm (e) groups. There was no significant difference between the prediction and clinical data for (d) (p=0.20) or (e) (p=0.69).

Using our mathematical framework and these estimated rates, we then calculated the probability (as a function of tumor burden) that chemo-resistant cells are present at diagnosis (Figure S3a). Consistent with clinical and theoretical studies of other malignancies^37-39,41-45^, our model predicted that at least some chemo-resistant cells are always present at the start of therapy for HGSC. We varied the values for each model parameter over a large range to test their influence on the calculated number of resistant cells at diagnosis: within the ranges tested, this number almost always exceeds 10^3^ (Figures S3b-g).

### Model Validation

To assess the accuracy of our mathematical framework and the validity of the estimated parameter values, we analyzed the model-predicted distribution of overall survival using data from several test sets (Figure 3). First, we predicted the survival of patients in Validation Set 1 (Figures 3a-b, red curves), by adjusting the value of *M*. Notably, our model recapitulated the clinical observation that residual tumor size is significantly associated with survival in patients undergoing PDS, but not NACT (Figures 3a-b). Comparison of the observed (Figures 1c-d, red curves) and predicted (Figures 3a-b, red curves) distributions of patient survival revealed no significant differences (Figures 3c-d).

**Figure 3.**
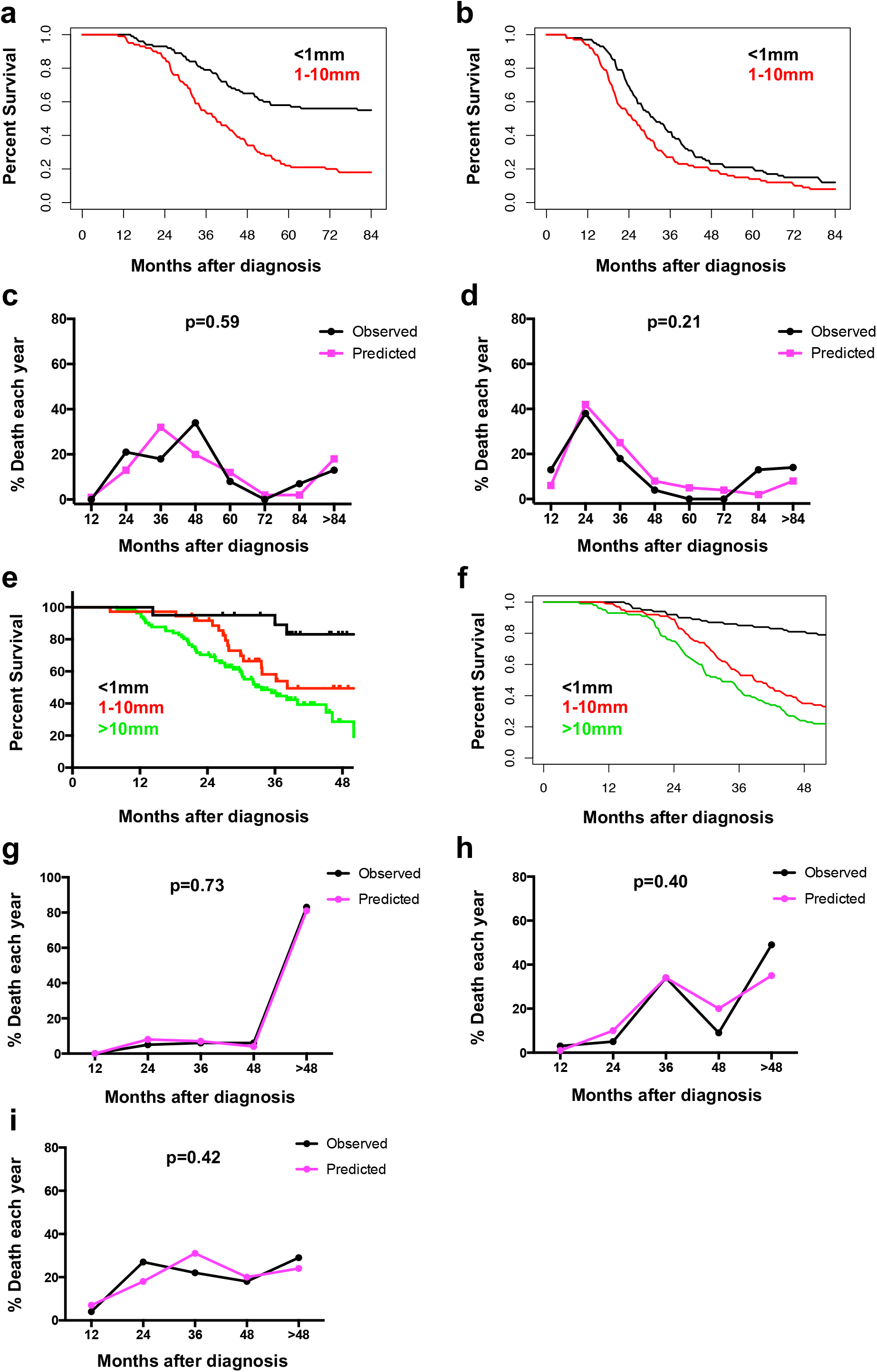
Evaluation of Model Predictions with Data from Validation Sets. (a-b) Predicted survival of patients from the UHN dataset treated with PDS (a) or NACT (*b*). Black curves represent the predicted overall survival curves for patients with <1mm residual tumor. Red curves represent the predicted overall survival curves for patients with 1-10mm residual tumor post-surgery. Note that the model predicts that post-surgery cancer size is critical for the outcome of PDS, but not NACT. (c-d) Comparison of predicted and observed survival for the PDS (c) and NACT (d) 1-10mm residual tumor groups. There was no significant difference between the predictions and clinical data for either (c) (p=0.59) or (d) (p=0.21). (e-f) Observed (e) and predicted (f) overall survival of patients in the NCIC cohort. Patients received PDS and had <1mm, 1-10mm, or >10mm residual tumor. (g-i) Predicted and observed survival of PDS-treated patients from the NCIC study with <1mm (g), 1-10mm (h), or >10mm (i) residual tumor. There was no significant difference between predictions and clinical data for (g) (p=0.73), (h) (p=0.40), or (i) (p=0.42). Parameter values were the same as in Figure 2c, except that *log_10_M* was set with mean 8 and s.d. 0.4 for the 1-10mm residual group, and mean 10 and s.d. 0.4 for the >10mm residual groups. Also, the gap between surgery and the start of chemotherapy was 1 month for the UHN patients and 0.8 month for the NCIC patients; UHN patients received 6 cycles of chemotherapy for first line PDS treatment, whereas NCIC patients received 8 cycles.

The data for the above validation exercise derived from different patients than those used for the Training Set, but all of these patients were treated over the same time period at the same institution (UHN). To more rigorously test the validity of our model, we analyzed independent patient data, derived from the NCIC cohort. As noted above, the treatment course of these patients also differed somewhat from that of the UHN patients, enabling an even better test of the general applicability of our framework. We incorporated these differences into the model and predicted the survival of patients in the NCIC trial. Again, the model predictions fit very well with clinical observations, and recapitulated the theme that PDS with minimal residual tumor results in the best outcome (Figures 3e-i).

### Predicted Outcome of PDS and NACT in Patients with Identical Tumor Burden

Confident in the predictive ability of our mathematical framework, we modeled the expected clinical outcome of PDS and NACT in patients with the same initial tumor burden by imposing the same distribution of *M_1_* on both groups. Our model predicts that PDS patients should survive longer than NACT patients when controlled for residual post- surgery tumor mass. As residual tumor increases, however, the predicted survival advantage of PDS shrinks (Figures 4a-b). To investigate the underlying reason(s) for these predictions, we explored the predicted dynamics of chemo-sensitive (green) and chemo- resistant (blue) HGSC cells in patients undergoing treatment by PDS or NACT (Figures 4c-d). Our model predicts that, at diagnosis, a typical HGSC patient has low numbers of chemo-resistant cells. In women who undergo PDS, debulking surgery (S in Figure 4c) dramatically reduces the number of chemo-sensitive and chemo-resistant cancer cells, because these cells appear identical to the surgeon and therefore have an equal likelihood of removal. Depending on the (stochastic) distribution of chemo-resistant cells within the abdominal cavity of the HGSC patient, all chemo-resistant cells present at diagnosis might have been eliminated by complete debulking, with the residual chemo-resistant cell number following a Poisson distribution. Follow-up chemotherapy (C in Figure 4c) can then reduce the remaining chemo-sensitive cells to very low numbers or even eradicate them.

**Figure 4.**
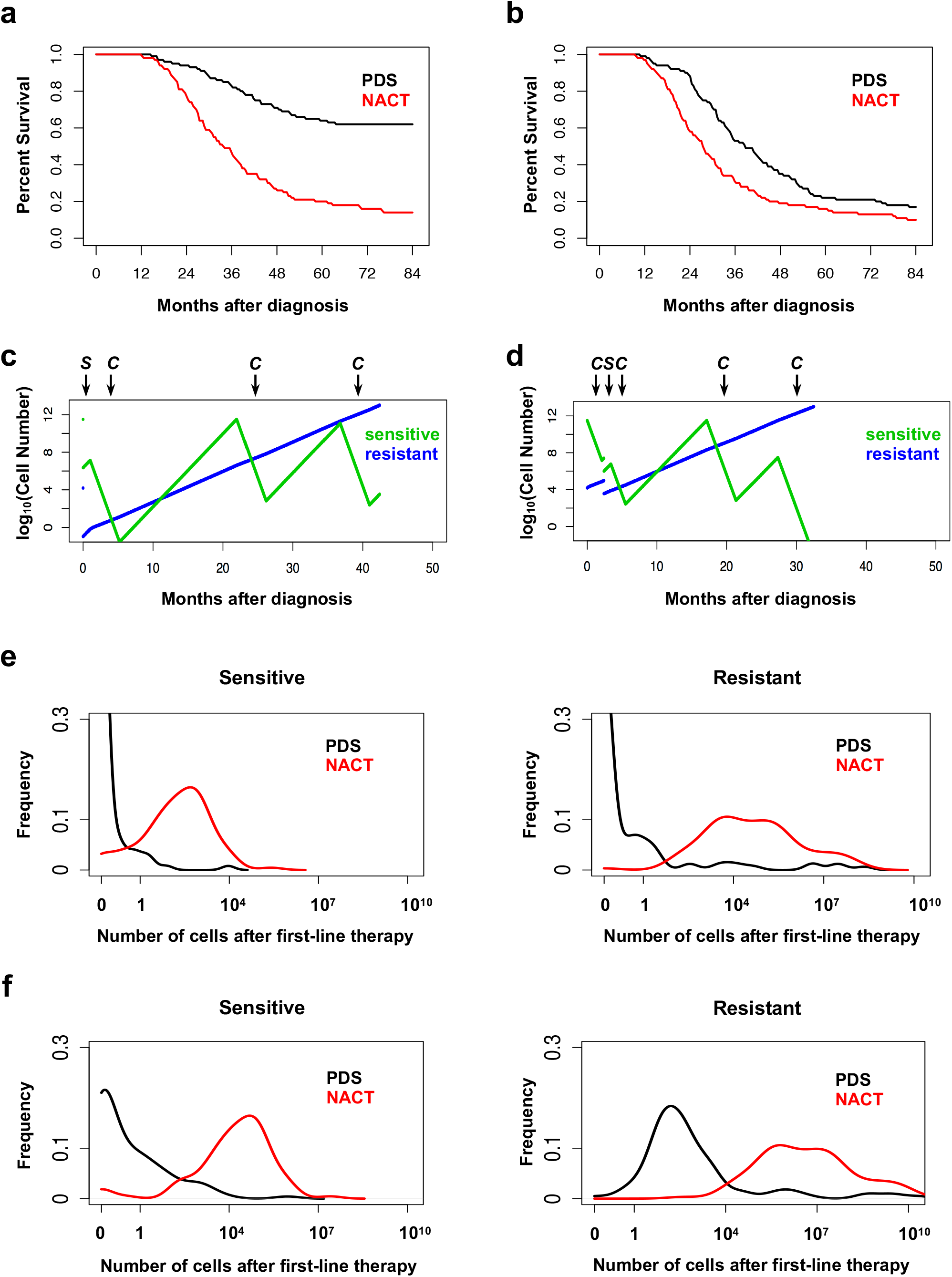
Predicted Outcome of PDS and NACT Patients with Same Initial Tumor Burden. (a-b) Predicted survival of patients undergoing PDS (black curves) or NACT (red curves). All patients received 6 cycles of chemotherapy. Panel (a) shows the results for <1mm residual tumor group, and panel (*b*) shows the result for 1-10mm residual tumor group. Parameter values were as in Figure 2c, except that *log_10_M_1_* was set at the same value for the PDS and NACT groups, with mean 11.5 and s.d. 0.4. (c-d) Simulation of representative progression dynamics for chemo-sensitive (green curves) and chemo-resistant (blue curves) cancer cells in a patient undergoing PDS (c) or NACT (d) treatment with optimal debulking. Treatment order is shown at the top of each plot; “S” indicates “surgery”, and “C” indicates “chemotherapy”. (e-f) Distribution of number of chemo-sensitive and chemo-resistant cells for the PDS (black curves) and NACT (red curves) groups, with <1mm (e) or 1-10mm (f) residual tumor.

By contrast, with NACT, neo-adjuvant chemotherapy (C) dramatically enriches for chemo-resistant cells, while killing the sensitive cells; consequently, chemo-resistant cells comprise a large proportion of total tumor cells at surgery (S). Because chemo-sensitive cells are largely depleted by the neo-adjuvant chemotherapy, the amount of residual tumor visible to the surgeon is reduced substantially. Consequently, it is virtually impossible for interval debulking surgery to fully deplete the chemo-resistant cells (Figure 4d). We propose that the relative inability of NACT to deplete chemo-resistant cells underlies the difference in outcome between patients treated with PDS and NACT. Importantly, this conclusion is based on intrinsic properties of the dynamics of cancer proliferation, survival, and death.

We then explored why the predicted superiority of PDS over NACT depends on residual tumor burden post-surgery. By examining the expected distribution of chemo- sensitive and –resistant cell numbers after first-line therapy, we found that PDS with <1mm residual tumor has the potential to deplete all cancer cells in a significant proportion of HGSC patients (Figure 4e). This finding can account for the considerable survival difference between PDS and NACT with <1mm residual tumor. By contrast, with >1mm residual tumor, neither PDS nor NACT depletes all malignant cells, even though fewer tumor cells are predicted to remain after PDS (Figure 4f). Consequently, almost all patients with >1mm residual tumor are predicted to relapse and eventually die because of the inability of current agents to kill chemo-resistant cells. This analysis explains why PDS is superior to NACT when complete debulking can be achieved and why residual tumor mass is a key determinant of survival after PDS but not NACT (Figures 3a-b).

### Test the robustness and generalizability of model prediction

To test the robustness of our predictions and quantitatively examine the influence of each factor on survival, we varied the values of every parameter in our model over a large range. Our conclusions on the relative efficacy of PDS and NACT treatment with complete debulking hold true throughout, although the magnitude of the predicted difference can vary (Figure S4). Systematic modulation of the model parameters also enabled us to assess the contribution of each factor to patient survival, and revealed several interesting features. For example, we found that faster growth of cancer cells does not necessarily lead to worse survival. In particular, although elevated growth rate of chemo- resistant cells in the absence of chemotherapy leads to worse survival, faster-growing, chemo-sensitive cells can sometimes lead to better survival (Figures S4a-d). This result can be attributed to a lower percentage of chemo-resistant cells at diagnosis when chemo- sensitive cells proliferate faster. However, if the proliferation rate of chemo-sensitive cells is too high (e.g., when *r* = 3), new chemo-resistant cells might be generated between the time of surgery and adjuvant chemotherapy, resulting in worse survival (Figure S4c). We also inferred that the relative importance of chemo-sensitive and chemo-resistant cells in influencing survival might differ at different stages along the clinical course. During treatment-free periods, the growth rate of chemo-resistant cells could play a more dominant role in influencing patient survival (Figures S4a-d), because they underlie ultimate treatment failure. However, during periods of chemotherapy, the growth rate (or depletion rate) of chemo-sensitive cells might play a more dominant role (Figures S4e-h), because the depletion rate of chemo-sensitive cells at this stage determines whether they can be completely eradicated by chemotherapy. By contrast, chemo-resistant cells likely will endure. As a result, drug choice and dose, which likely influences the efficiency of elimination of chemo-sensitive cells, is predicted to be a critical factor in treatment outcome. Some molecular subgroups of HGSC (e.g., *CCNE1* -amplified tumors) are highly resistant to current chemotherapy. Our model predicts that such patients would be refractory to PDS, even if complete debulking is achieved (Figures S4e-f, when *r’* is high or *d’* is low). Residual cancer cell abundance after tumor resection can dramatically influence patient survival, especially in the PDS group (Figure S4i), suggesting that primary cytoreductive surgery should aim for complete removal of cancer cells even though chemotherapy will usually follow. Tumor size at diagnosis can be a critical factor determining whether a patient can be cured (Figure S4j), and will be further addressed below. By contrast, varying the parameter value of tumor size at patient death did not influence the length of survival (Figure S4k).

To further test the general applicability of our model, we tested whether it could explain the similar outcome of patients treated with PDS and NACT in prospective clinical studies, such as the CHORUS trial^28^, using their patient data. Patients in this study had remarkably shorter overall survival than most other study cohorts. We reasoned that this might be attributable to more severe disease at diagnosis in patients in this cohort, such as higher tumor burden and/or more severe systemic metastasis. We therefore re-trained the parameters *M_1_* and *ε* for patients in the CHORUS study, only using the data from PDS or NACT 1-9mm groups in this cohort. Indeed, the model predicted higher values for both parameters (median of *M_1_* = 10^12.2^ and *ε* = 10^-29^) (Figure S5a). Interestingly, when using these parameter values to predict the survival of PDS and NACT <1mm groups, the model predicted no significant difference between the two regimens, consistent with the trend published in the original study^28^ (Figure S5b). These results indicate that the contradiction between existing retrospective studies and prospective trials may have derived from the differences in the disease severity at diagnosis of patients in each cohort. It further suggests that disease burden information is a critical factor in the choosing PDS or NACT, and also reflects the need for personalized treatment even when only the temporal order of treatment is at stake.

We also considered two alternative scenarios: (1) that heterogeneous populations of (variably) chemo-resistant cells exist in the same patient (Figures S6a-c); or (2) that HGSC initiates from an intrinsically chemo-resistant “cancer stem cell”, which differentiates into chemo-sensitive “tumor progenitor cells” (Figures S6d-f). Either of these scenarios results in the same conclusions as the original model.

### Modeling Alternative Treatment Regimens

We next utilized our model to predict the effects of altering current treatment regimens. For both PDS and NACT, adjuvant chemotherapy typically begins 4-5 weeks post-surgery, an interval chosen to allow patients to recover from their typically aggressive surgical treatment. The length of the post-surgical chemotherapy delay varies between centers and among physicians, but its influence on treatment outcome has not been studied carefully. We varied the length of treatment delay in our model, and tested the predicted effects on patient survival. For PDS with <1mm residual tumor, earlier initiation of chemotherapy should prolong survival, whereas longer treatment delay worsens outcome (Figure 5a, *p*<0.0001). For PDS with >1mm residual tumor or for NACT with any amount of residual tumor, the length of treatment delay (within the same range) is predicted to have little effect on outcome (Figures 5b-d). These differences arise primarily because upfront surgery that results in <1mm residual tumor, followed by chemotherapy, potentially can deplete all tumor cells when treatment delay is minimized (Figure 4e). The probability of depletion decreases with longer delay, largely because chemo-resistant cells can arise during the gap between upfront surgery and adjuvant chemotherapy (Figure S7a). By contrast, PDS with >1mm residual tumor or NACT is unlikely to deplete all cancer cells (Figures 4e-f and S7b-d), irrespective of treatment delay.

**Figure 5.**
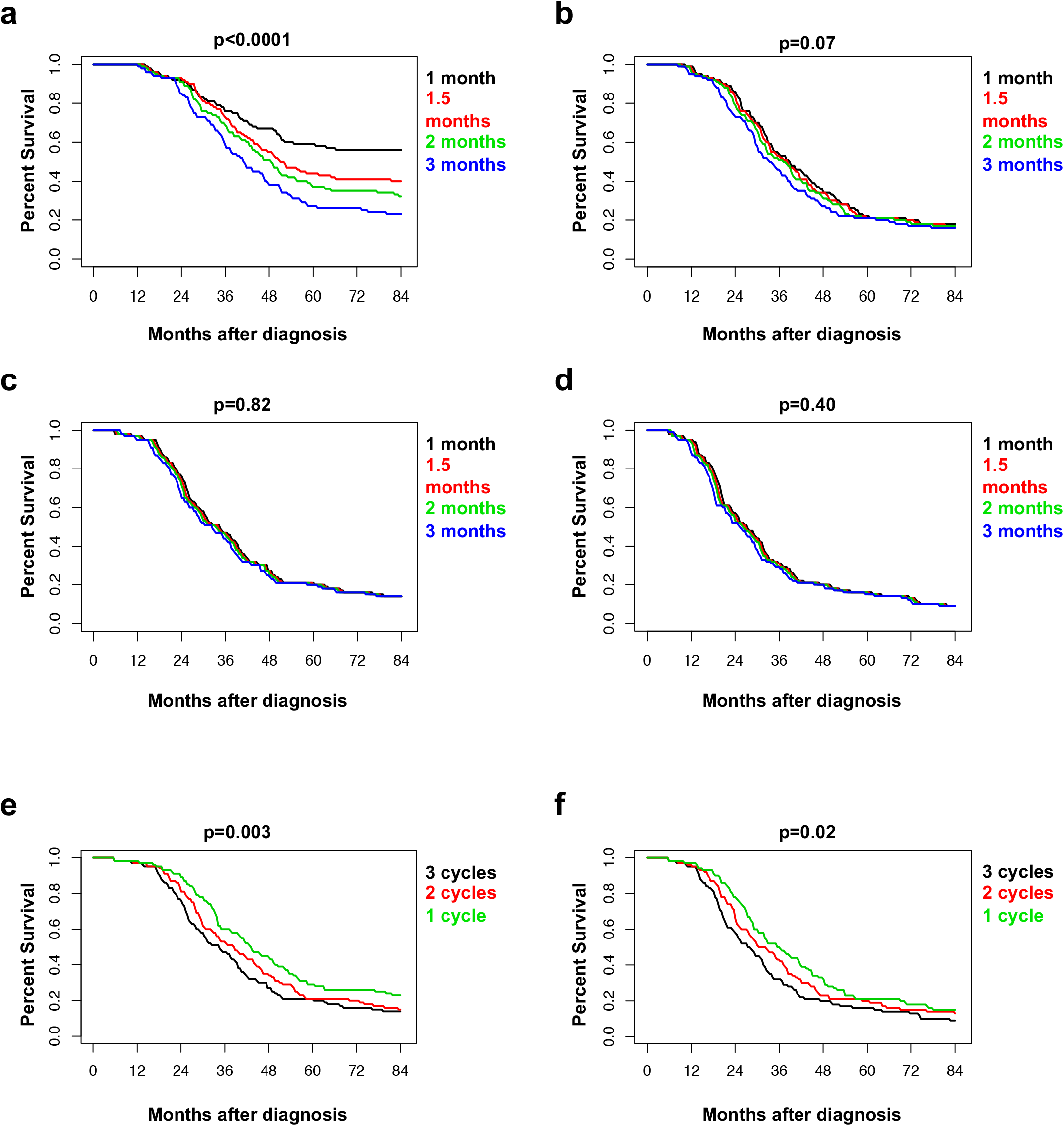
Predicted Outcomes of Alternative Treatment Strategies. (a-d) Predicted survival of patients receiving PDS (a-b) or NACT (c-d) with different lengths of delay between debulking surgery and adjuvant chemotherapy. Panels (a) and (c) show expected results with <1mm residual tumor after surgery; panels (*b*) and (d) show predicted results with 1-10mm residual tumor. (e-f) Predicted overall survival of patients receiving NACT with different cycles of neo-adjuvant chemotherapy before and after surgery, with options of: 3 neo-adjuvant and 3 adjuvant cycles (black curves), 2 neo-adjuvant and 4 adjuvant cycles (red curves), or 1 neo-adjuvant and 5 adjuvant cycles (green curves). Panel (e) shows the expected results with <1mm residual tumor; panel (f) shows expected results with 1-10mm residual tumor. Parameter values are the same as in Figure 2c.

Our model predicts that neo-adjuvant chemotherapy enriches for chemo-resistant cells and thus increases the percentage of chemo-resistant tumor cells remaining post-surgery in patients treated with NACT. Conceivably, reducing the number of cycles of neo-adjuvant chemotherapy could attenuate the enrichment for chemo-resistant cells, and if the same number of residual cancer cells remain after surgery, less neo-adjuvant chemotherapy might prolong survival. Indeed, our model predicts that patients undergoing NACT with <1mm or >1mm residual tumor could benefit slightly from reducing the number of pre-surgery chemotherapy cycles (Figures 5e-f). This small predicted improvement arises primarily because of more efficient reduction of the number of chemo-resistant cells at surgery. The benefit is limited, however, because for most patients, chemo-resistant cells are still unlikely to be eradicated after their enrichment during the neo-adjuvant chemotherapy period (Figures S7e-f).

### Predicted Effects of Earlier Diagnosis on Survival

We also utilized our model to evaluate the potential benefits of earlier diagnosis. We first modeled the effects of diagnosing relapsed HGSC at the earliest possible time enabled by the currently used clinical test, CA-125 detection^46^. Contrary to the intuitive notion that earlier diagnosis of recurrence should be advantageous, we find that CA-125- based earlier diagnosis is not expected to improve survival (Figures 6a-b, red curves). We also asked if detecting recurrence earlier than is possible with CA-125 monitoring would be advantageous. Yet even with a lead time ~10^4^ greater than that required for physical symptoms to appear, the current upper limit of sensitivity for ctDNA-based diagnosis of ovarian cancer^33^, our model predicts no advantage in patient survival with earlier detection of recurrence (Figures 6a-b, green curves).

**Figure 6.**
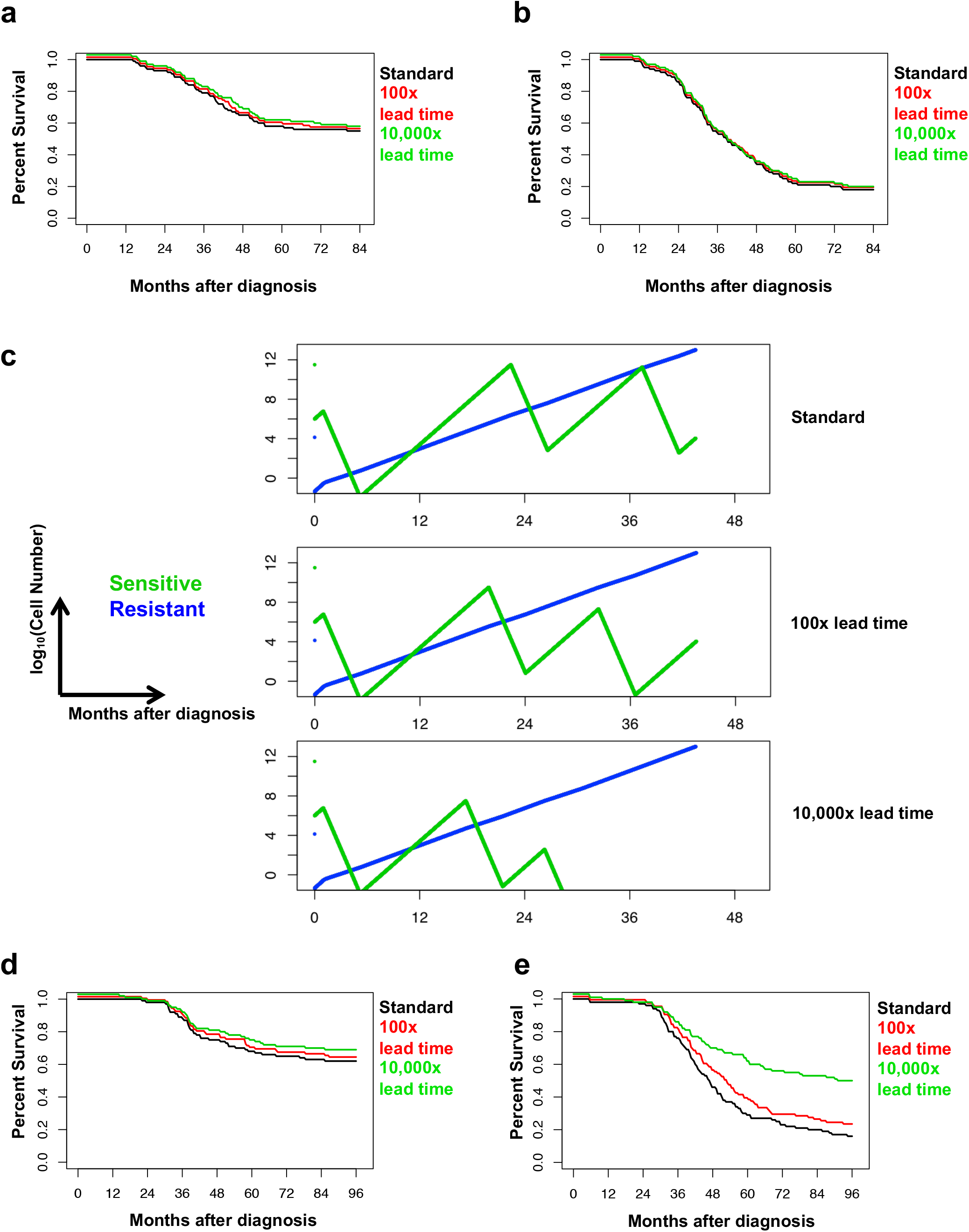
Predicted Effects of Earlier Diagnosis and Treatment of Relapsed Cancer. (a-b) Predicted survival of patients with relapsed cancer detected at different degrees of sensitivity. Predictions were stratified based on the first-line therapy performed, including (a) PDS or (b) NACT with <1mm residual tumor. “Standard” (black curves) represents diagnosis based on physical symptoms, when the number of cancer cells is comparable to *M_1_*; “100x lead time” (red curves) and “10,000x lead time” (green curves) represent earlier treatment of recurrent disease with the number of cancer cells at diagnosis 1% or 0.01% that of *M_1_*, respectively. (c) Simulation of representative growth dynamics for chemo-sensitive (green curves) and chemo-resistant (blue curves) cancer cells in a patient treated by PDS with optimal debulking, with relapsed cancer diagnosed at different degrees of sensitivity. Earlier treatment of relapsed cancer can more effectively deplete chemo-sensitive cells, but does not effectively change the trajectory of chemo-resistant cells, which are the ultimate cause of patient death. Parameter values are the same as in Figure 2c. (d-e) Predicted survival of patients with relapsed cancer detected at different degrees of sensitivity and treated with a different second-line chemotherapy. Predictions were stratified based on the first-line therapy performed, including (d) PDS or (e) NACT with <1mm residual tumor. “Standard” (black curves) represents diagnosis based on physical symptoms, when the number of cancer cells is comparable to *M_1_;* “100x lead time” (red curves) and “10,000x lead time” (green curves) represent earlier treatment of recurrent disease with the number of cancer cells at diagnosis 1% or 0.01% that of *M_1_*, respectively.

We explored the reason for this lack of survival advantage by modeling the theoretical numbers of chemo-sensitive and –resistant cells in a virtual patient whose recurrence is diagnosed with increasing levels of sensitivity (Figure 6c). Although earlier diagnosis, followed by prompt re-institution of chemotherapy, can better deplete chemo- sensitive cells, it barely affects the chemo-resistant cells that have been enriched by first-line therapy, which ultimately expand and cause patient death. Therefore, earlier diagnosis is unlikely to improve survival when applied to relapsed cancers treated with standard cytotoxic chemotherapy regimens. Based on this result, we asked if treating relapsed tumors using a hypothetical drug with similar efficacy but distinct resistance mechanism from platinum/paclitaxel would potentiate the benefit of earlier diagnosis of relapsed HGSC. Indeed, our model predicts that earlier diagnosis of relapsed cancer can be beneficial, but only with alternative second-line therapy (Figure 6d-e). The magnitude of the advantage of earlier, over currently standard, diagnosis at relapse largely depends on the probability that earlier, but not later, intervention at relapse can be curative.

Finally, we used our model to explore the potential benefit of earlier diagnosis of treatment-naïve tumors at first detection in the clinic. We reasoned that as HGSC deposits usually get larger and/or more disseminated if left untreated, then earlier upfront diagnosis would likely identify smaller, probably less disseminated, tumors, potentially increasing the chances of complete debulking. We therefore focused our comparison on the predicted effects in patients with <1mm residual tumor. Our analysis argues that earlier diagnosis of treatment-naïve cancer, with concomitant prompt intervention, can improve patient survival compared to regular diagnosis when controlled for residual cancer cell number post-surgery (Figure 7). For PDS with complete debulking, the predicted survival benefit can be dramatic (Figures 7a, 7c, and 7e), primarily because lower volume and less diffuse tumor at presentation can increase the likelihood of disease eradication (Figures S8a, S8c, and S8e). By contrast, for NACT with complete debulking, the predicted benefit is rather limited, and the difference is detectable only if lead time is sufficient (Figures 7b, 7d, and 7f). In that case, chemo-resistant cells might not yet have arisen, and chemotherapy alone might be sufficient to eradicate disease (Figures S8b, S8d, and S8f).

**Figure 7.**
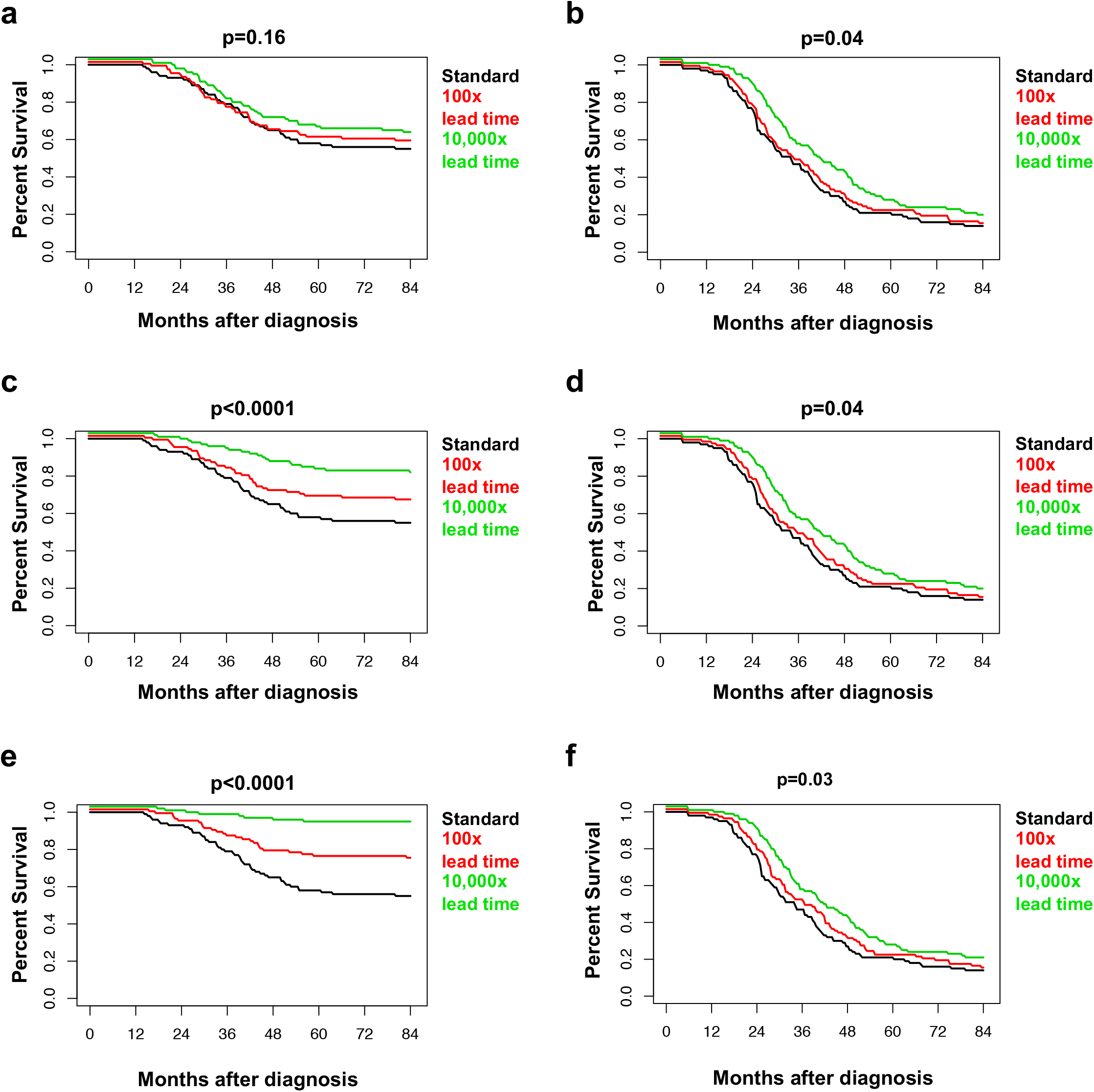
Predicted Advantage of Earlier Diagnosis and Treatment of Naïve Cancer. Predicted survival of patients diagnosed with treatment-naive cancer at different degrees of sensitivity. Predictions were stratified based on the eventual first-line therapy received, including PDS (a, c, e) and NACT (b, d, f) with <1mm residual tumor. “Standard” (black curves) indicates diagnosis based on physical symptoms, when the number of cancer cells is equal to *M_1_* “100x lead time” (red curves) and “10,000x lead time” (green curves) represent earlier treatment of naïve cancer, with the number of cancer cells at diagnosis 1% or 0.01% that of *M_1_*, respectively. Because earlier detection is likely to be associated with less cancer dissemination, the numbers of residual cancer cells post-surgery might be smaller. We tested scenarios wherein 100x earlier detection is associated with 100% (a-b), 50% (c-d), or 20% (e-f) the disease diffusion of Standard detection, by adjusting the number of residual tumor cells for the “100x lead time” group to 1, 1/2, or 1/5 of *M* for “Standard” group, respectively. Other parameter values are the same as in Figure 2c.

## Discussion

Mathematical modeling has demonstrated potential in the systematic and quantitative assessment of various treatments^44,47,48^. If the outcomes of different strategies could be modeled accurately *a priori*, clinical trials could focus on therapeutic combinations that are most likely to succeed, and improvements in patient outcomes could be accelerated. The potential benefits of modeling are particularly important for diseases with relatively limited patient populations, in which it is not possible to do multiple clinical trials in parallel. Predictive models also can suggest when specific clinical controversies merit re-examination in the controlled trial setting. Our mathematical model of HGSC defines factors that affect the evolution of chemotherapy resistance and can predict patient survival based on the growth dynamics of chemo-sensitive and –resistant cells, whether stochastic or hierarchical models of tumor initiation are assumed. Furthermore, the results of our analyses have important implications for HGSC therapy and screening.

We populated our model with clinical data from ~300 patients receiving PDS or NACT. After estimating the rates of tumor cell proliferation and conversion to chemo- resistance, we concluded that most HGSC patients probably harbor chemo-resistant cancer cells at diagnosis. Our outcome predictions closely match patient data from multiple sources and support clinical observations that: (1) PDS that leaves minimal residual tumor is the optimal treatment strategy; (2) residual tumor size is a critical determinant of survival in patients undergoing PDS, but not NACT; (3) earlier diagnosis of relapsed cancer does not - and cannot - lead to better survival with current therapies; and (4) earlier diagnosis of primary (treatment-naïve) HGSC could dramatically improve outcomes from this devastating disease, if PDS with complete debulking is feasible.

Our model shows clearly that, when controlled for residual cancer size after surgery, PDS with complete debulking should lead to a better outcome than NACT with complete debulking. We infer that the reason for the superior predicted outcome of PDS is that upfront surgery can deplete minority chemo-resistant cells more effectively, leaving adjuvant chemotherapy to eradicate residual chemo-sensitive cells. If debulking surgery removes all chemo-resistant cells, the PDS regimen can be curative. By contrast, cure is unlikely after NACT for at least two reasons. First, neo-adjuvant chemotherapy enriches for, and allows the continued expansion of, chemo-resistant cells. Second, by depleting bulk chemo-sensitive cells, chemotherapy removes tumor mass that can mark the location of chemo-resistant cells interspersed in metastatic deposits throughout the peritoneum. We suggest that removing these “sentinel” chemo-sensitive cells renders interval surgery less effective than upfront surgery in depleting chemo-resistant cells, which are the cells that eventually cause death. Put another way, with existing agents, the only way to achieve cure, and the best way to promote long-term survival, is to remove all, or the vast majority of, chemo-resistant cells. At present, the only way to achieve this goal is by surgically removing such cells, which is much more likely with the PDS regime. Of course, the development of drugs that can target these resistant cells might alter these conclusions^17,22,49^.

Previous clinical studies differed over whether PDS or NACT is superior. A retrospective analysis of patients treated between 1980 and 1997 found no difference in outcome between PDS and NACT^26^. By contrast, several meta-analyses indicated that NACT is associated with worse prognosis^21,50^. This controversy occasioned two large, controlled clinical trials to compare PDS and NACT^23,28^, which found no significant difference in patient survival between PDS and NACT. Three major weaknesses limit the strength of these trials’ conclusions: (1) They only recruited patients with “extensive” HGSC, which might explain the notably worse outcome of patients in this study, especially in PDS <1mm group, compared with multiple other reports^19,20,24,51^. Our computational model predicts that elevated upfront tumor burden can lead to worse overall survival and obscure the difference in outcome between PDS and NACT, even when complete debulking is performed (Figure S4j). In particular, differences in the magnitude of *ε*, the “inaccessible proportion”, alone might contribute to the lack of difference between PDS and NACT in these trials. (2) In the EORTC trial, there was marked inconsistency between the participating institutions in percentage of patients successfully undergoing optimal debulking in the PDS group, with 63% for one center in a single country and <12% for the other 6 countries involved^23,52^. Our model predicts that residual tumor size is a critical factor for patient survival, especially in the PDS group, and increased abundance of residual cancer cells can decrease the advantage of PDS (Figure S4i). (3) In the CHORUS trial, many patients (30-40%) received carboplatin monotherapy, instead of combination carboplatin and paclitaxel^28^. Previous studies demonstrated that platinum-based combination therapy in ovarian cancer leads to better survival than platinum therapy alone^53^, likely by more effectively depleting chemo-sensitive cells or by killing some platinum-resistant cells. Our model predicts that the efficiency of depletion of chemo- sensitive cells during chemotherapy is a critical factor for patient survival (Figure S4e-f). Therefore, the model’s predictions might explain the different findings from existing clinical studies.

A previous mathematical modeling study, based on a Gompertzian growth model, argued that NACT should be superior to PDS^54^. However, that study did not account for the different dynamics of chemo-sensitive and –resistant cells, and assumed equal efficiency of surgical depletion of large and small tumors, which ignores the intrinsic limitations of surgery. Such assumptions can lead to serious errors in modeling HGSC. For example, although aggressive surgery might reduce a tumor containing >10^11^ cells to a mass of <10^7^ cells, a tumor containing <10^6^ cells is unlikely to be visible during surgery, making it highly unlikely that such a tumor can be surgically reduced to <10^2^ cells. Notably, the conclusions of our study still hold if our model is implemented with Gompertzian growth assumptions (data not shown).

Similarly, a study of the optimal order of surgery and chemotherapy in pancreatic cancer concluded that neo-adjuvant chemotherapy should be superior to upfront surgery^36^. However, the biology and chemo-responsiveness of pancreatic cancer and HGSC differ substantially: whereas HGSC is usually quite chemo-responsive, pancreatic cancer is notoriously chemo-resistant. The differential influence on tumor visibility following chemotherapy likely underlies the different predictions in the two diseases. Nevertheless, the difference between our conclusions and those of Haeno *et al*. argue for caution in extrapolating their conclusions to other types of cancer.

A limitation of our study is that some practical considerations of treatment are beyond the scope of computational modeling. For example, in many patients (e.g., the infirm), NACT is preferred simply because of the potential risks of this large operation, which include bowel perforation, uncontrolled bleeding and/or the stresses of prolonged surgery/anesthesia. In such scenarios, treatment choice depends primarily on technical feasibility.

In addition to fitting existing clinical studies comparing PDS and NACT, our model enables quantitative analysis of the dependence of treatment outcome on various factors under different scenarios, providing a powerful tool to assist clinical decision-making. For example, we found that the advantage of PDS over NACT diminishes with larger residual tumor post-surgery, more extensive metastases at unresectable locations, and/or a higher percentage of chemo-resistant cells at diagnosis (Figures 3a-b, and S4a-c). These features might help explain the similar overall survival between PDS and NACT groups in studies involving patients with more extensive upfront disease or less complete surgical removal^23,26^. Our analysis also argues that for PDS patients with <1mm residual tumor, adjuvant chemotherapy should start as early as possible to provide the best chance of curative outcome; indeed, we predict that differences of even a few weeks might dramatically alter the chance for curative outcome. Conversely, if complete debulking is not achieved at primary surgery, delaying chemotherapy is less likely to affect survival. These predictions are consistent with the results of a meta-analysis of clinical studies^55^, which found that each extra week of delay was associated with a significantly decreased overall survival in patients with PDS <1mm residual tumor, but not in patients with visible residual disease. We also predict that reducing the number of cycles of neo-adjuvant chemotherapy should result in slightly improved overall survival for NACT patients, a prediction that also is consistent with a meta-analysis of clinical data^50^. Taken together, these findings argue that even in patients too infirm to undergo immediate surgical debulking, the number of neo-adjuvant cycles before surgery should be minimized.

A final prediction of our model is that the effects of earlier diagnosis of HGSC should be quite different in relapsed versus treatment-naïve patients. For the former, earlier diagnosis is unlikely to improve overall survival, primarily because earlier treatment of recurrent tumors with existing agents cannot deplete chemo-resistant cells that have been enriched over the clinical course. This prediction matches very well with earlier clinical observations^31^. Two scenarios might alter this conclusion: (1) if effective chemotherapy with a resistance profile orthogonal to platinum/taxane-based therapy were to become available at relapse; or (2) if effective debulking could be achieved at relapse. Effective drugs against HGSC remain a major clinical limitation^56,57^. Our findings argue strongly for employing alternative agents as early as possible upon tumor relapse. Surgery is no longer typically performed on recurrent HGSC, and its efficacy could be limited by the proportion of cancer cells at unresectable locations at disease relapse. Nevertheless, our model calls for re-evaluation of feasibility and potential efficacy of secondary surgery for the benefit of earlier diagnosis. Consistent with our analysis, several retrospective studies report that secondary surgery achieving complete debulking can be beneficial for HGSC patients with platinum-sensitive tumors^58,59^.

By contrast, our model predicts that earlier diagnosis of treatment-naïve cancer could improve overall survival for at least two reasons: (1) chemo-resistant cells are usually not enriched prior to treatment and earlier intervention can reduce the likelihood that significant numbers will arise; and (2) earlier upfront surgery has a better chance of removing all chemo-resistant cells, assuming that tumor cells have not diffused throughout the peritoneal cavity or seeded unresectable locations. This trend is consistent with a recent clinical trial testing the benefit of earlier upfront diagnosis of HGSC, which found that women who were screened annually for serum CA125 levels had reduced mortality from ovarian cancer than those with no screening^60^. In that study, the survival improvement by CA125-based screening was significant but modest, and might be attributable to at least two factors: (1) the lead time of CA125-based screening is on average less than 1 year^61,62^, which is shorter than the screening interval, rendering some patients not diagnosed earlier; (2) early-diagnosed patients may receive either PDS or NACT treatment, and according to our prediction the use of NACT might counteract the benefit of earlier diagnosis. In addition, there might be two potential benefits of earlier diagnosis and treatment of naïve HGSC not accounted for in our model: (1) earlier diagnosis might increase the likelihood of complete debulking; and (2) patients considered treatable only by NACT with regular diagnosis might be eligible for PDS with earlier diagnosis.

In summary, our analyses suggest that future randomized clinical trials might consider: (1) the influence of the interval between primary debulking surgery and adjuvant chemotherapy on treatment outcome; (2) the association between the number of neo-adjuvant chemotherapy cycles and treatment outcome; and (3) the effects of alternative chemotherapy and/or complete secondary surgery on relapsed tumor, especially when coupled to earlier diagnosis. Our results also predict that whereas not all patients who can achieve complete cytoreduction will benefit from PDS over NACT (give the importance of, and difficulty in measuring, *ε*), there is a subset of patients whose only chance of long-term survival or cure is the former regime. Finally, the mathematical abstraction makes our framework potentially applicable to treatment of other malignancies with alternative parameter values.

## Materials and Methods

### Patient Information

Clinical data from 285 HGSC patients were obtained from patient records at University Health Network (148 patients) and from the National Cancer Institute Canada OV16 trial (137 patients). Medical record information, including date of diagnosis, age of patient, timing of treatment, extent of residual disease in diameter (<1mm, 1-10mm, or >10mm), CA125 levels along the treatment course, and survival data (updated in 2014 for UHN data and 2010 for NCIC data) were recorded. Institutional REB approval was obtained through University Health Network and a Data Sharing Agreement was concluded with National Cancer Institute Canada.

### Variables Used in Mathematical Model

We denote the following variables for the development of mathematical modeling: *r*, division rate of chemo-sensitive cells in the absence of chemotherapy; *d*, death rate of chemo-sensitive cells in the absence of chemotherapy; *a*, division rate of chemo-resistant cells in the absence of chemotherapy; *b*, death rate of chemo-resistant cells in the absence of chemotherapy; *r’*, division rate of chemo-sensitive cells in the presence of chemotherapy; *d’*, death rate of chemo-sensitive cells in the presence of chemotherapy; *a’*, division rate of chemo-resistant cells in the presence of chemotherapy; *b*’, death rate of chemo-resistant cells in the presence of chemotherapy; *M*, the number of residual cancer cells in the peritoneal cavity after surgery; *M_1_*, the total number of cancer cells at diagnosis; *M_2_*, the total number of cancer cells at death.

### Estimation of Parameter Values

Unless otherwise specified, clinically relevant parameter values were estimated and set as follows:

In the absence of chemotherapy, the proliferation rate of chemo-sensitive cells (r) was obtained from the normal distribution with mean 2 and standard deviation 0.8, based on a clinical study interrogating the doubling time of ovarian cancer cells by BrdU labeling^63^. Another study compared cancer cell proliferation rates in platinum responders and non-responders by thymidine labeling, and found that responders had significantly higher labeling index than patients with stable or progressive disease^64^. This analysis is consistent with several other studies that also found chemo-sensitive cells with proliferation advantage in the absence of therapy^65,66^. Accordingly, we set the proliferation rate of chemo-resistant cells (*a*) to a normal distribution with mean 0.84 and standard deviation 0.42. Death rates (*d* and *b*) were set as 10% of proliferation rates (*r* and *a)* respectively, which is within physiologically relevant range; in any case, varying this ratio over a large range does not affect the main conclusions of this paper.

During chemotherapy, the proliferation rate of chemo-sensitive cells (*r*’) was set to 10% of that of randomly growing cells (*r*); the death rate of chemo-sensitive cells (*d*’) was set to a normal distribution with mean 4.9 and standard deviation 1. These values were set to match the clinical observation that chemo-responsive relapse occurs on average ~11 months after 6 cycles of chemotherapy, indicating that cytotoxic reduction of chemo-sensitive cells by 6 cycles of chemotherapy corresponds to ~11 months of proliferation. Our inference is consistent with published efficacy of chemotherapy in HGSC^25,67^. The proliferation rate of chemo-resistant cells during chemotherapy *(a’)* was set to the same as that during random growth (*a*); the death rate of chemo-resistant cells during chemotherapy (*b*’) was set to twice as that during randomly growing (*b*), or 20% of *a’*, to reflect a modest effect of chemotherapy on chemo-resistant cells. Conversion rate during chemotherapy (*u*’) was set as 10 times of that during randomly growing (*u*), to reflect the DNA-damaging effect of platinum-based chemotherapy.

The parameters reflecting cancer cell numbers (*M*, *M_1_, M_2_)* were obtained from normal distributions in base 10 logarithmic scale. The number of residual cancer cells immediately post-surgery *(M)* was set to mean 6 and standard deviation 0.4 on the log scale for <1mm residual tumor, mean of 8 on the log scale for 1-10mm residual cancer, and mean of 10 on the log scale for >10mm residual cancer. The number of cancer cells at diagnosis *(M_1_)* was initially set at different values in patients receiving PDS and NACT, to reflect the clinical observation that patients with NACT tend to have more extensive disease at diagnosis^24^. *M_1_* for PDS was set at mean 11.5 and standard deviation 0.4 on the log scale, and *M_1_* for NACT was set at mean 12 and standard deviation 0.4; these values result in NACT patients starting with >3x the cancer burden in the model than those receiving PDS. Moreover, we varied the ratio of tumor burden at diagnosis in NACT vs PDS patients from 1 to 10^1.5^, and the main conclusions hold over the entire range. *M_2_* was set to mean 13 and standard deviation 0.4 on the log scale.

We varied the values for all of the above parameters to test the robustness of our conclusions. We found that the main conclusions hold true, though the magnitude of the differences between PDS and NACT treatment outcomes may vary.

### Mathematical Deduction of the Expected Number of Chemo-resistant Cells at Diagnosis

We deduced the expected the number of chemo-resistant cells at diagnosis based on previous studies^35,36^. We first calculated the probability that chemo-resistant cells exist at diagnosis *(P_d_)*, and then calculated the expected number of chemo-resistant cells at diagnosis (*Y_d_*).

To calculate *P_d_*, we summed the probabilities that the first successful lineage of chemo-resistant cells arises when there are 1, 2, 3, … *M_1-_1* chemo-sensitive cells. *P(x)* denotes the probability that the first lineage arises when there are *x* chemo-sensitive cells. *P(x)* can be expressed as the joint probability that no successful chemo-resistant lineage arises at 1, 2, 3, … *x*-1 chemo-sensitive cells, and that a successful lineage arises at exactly *x* chemo-sensitive cells. An expected 1/(1 − *d/r)* divisions are needed for an effective increase of 1 chemo-sensitive cell, and during these divisions an expected *u/(1 − d/r*) chemo-resistant cells are generated, among which a proportion (1 − *b/a)* will successfully persist. Assuming that the number of surviving chemo-resistant cells generated by each division follows a Poisson distribution with mean *(1 − b/a)u/(1 − d/r)^45^*, we derive *P(x):*

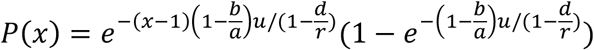

Thus the probability that chemo-resistant cells exist at diagnosis, *P_d_*, can be written as the sum of *P(x):*

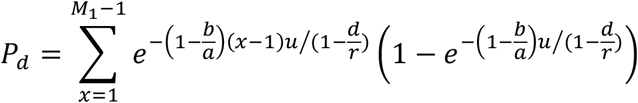

If we denote the time between the emergence of a successful chemo-resistant cell and diagnosis as *τ_x_*, then the expected number of chemo-resistant cells at diagnosis in the patients who have them can be expressed as:

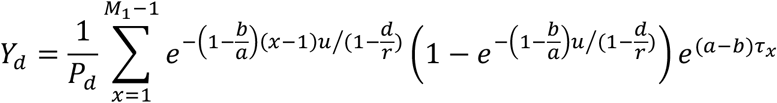

The amount of time *τ_x_* satisfies:

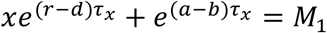

### Simulation of the Dynamics of Cancer Cell Number after Diagnosis

As the numbers of chemo-sensitive (denoted by *X)* and –resistant (denoted by *Y)* cells at diagnosis are expected to be large, we approximated their dynamics after diagnosis with a deterministic model, simulating the effects of random growth, chemotherapy, and surgery (cancer cell numbers before and after surgery are denoted as *X_before_, Y_before_*, and *X_after_, Y_after_*, respectively):

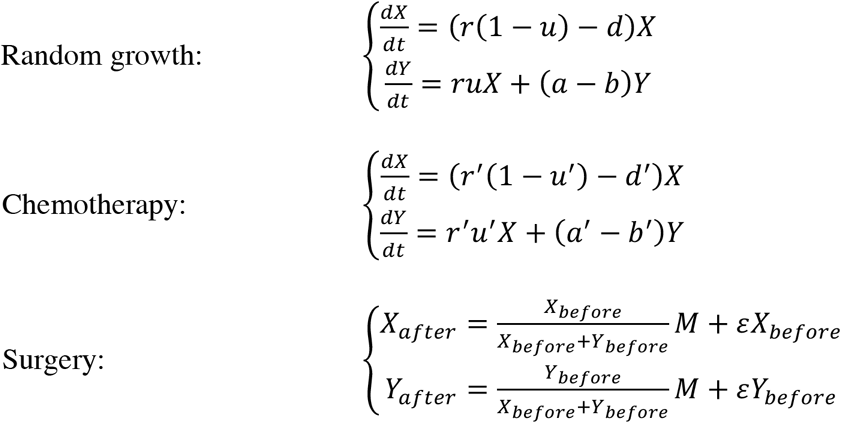

Treatment is adjudged a “failure” if the number of cancer cells post-treatment exceeds that pre-treatment. In the case of treatment failure, our model simulates random growth of cancer cells to the total number of *M_2_*, which marks patient death.

### Computational Estimation of Patient Survival

We applied the following workflow to simulate clinical treatment of HGSC. Overall survival is monitored and recorded along the workflow:

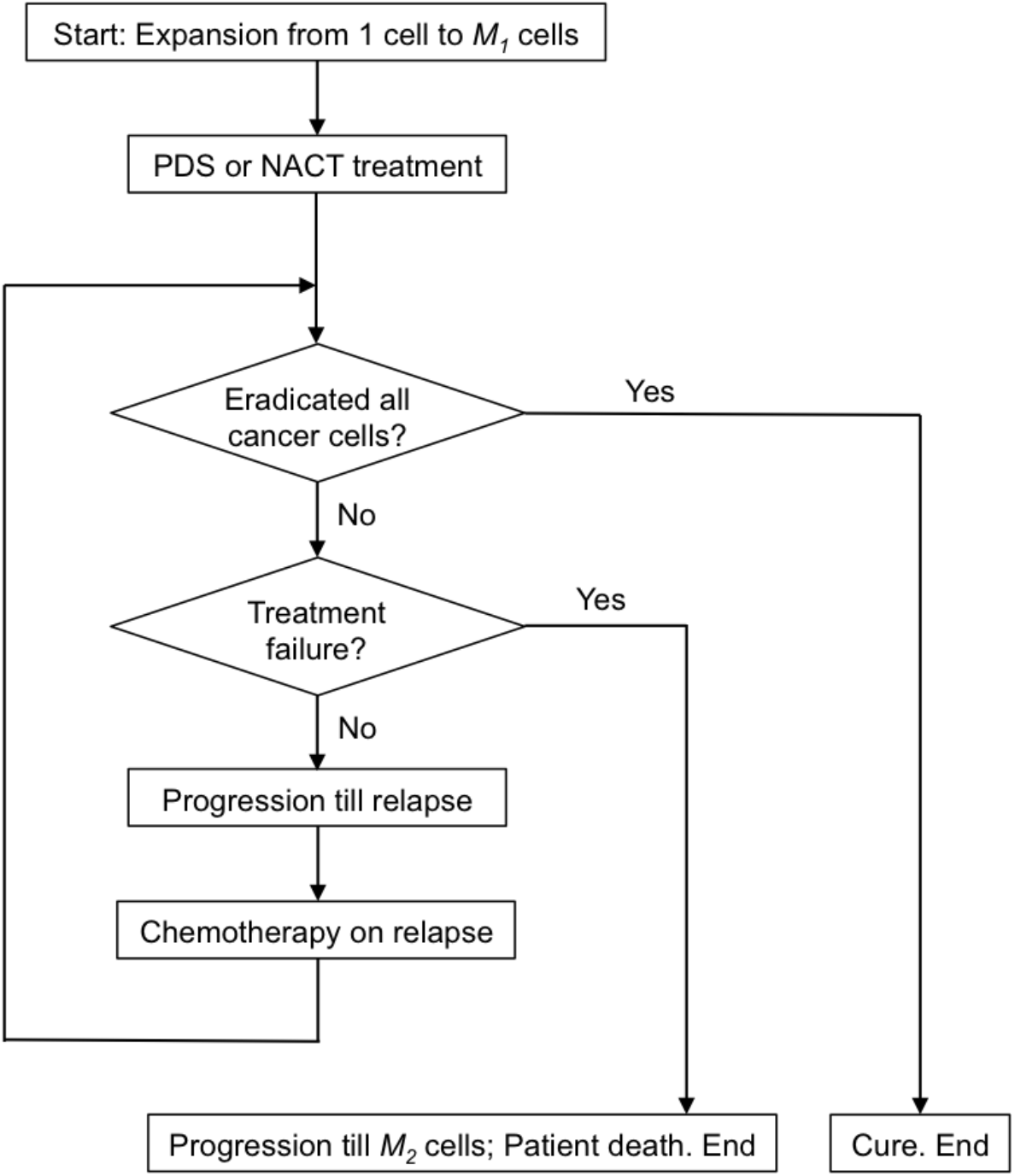

### Assessment of Prediction Robustness Regarding PDS vs. NACT Regimen

To assess the robustness of the model predictions of outcome following PDS vs. NACT, we varied *u*, *ε*, and each of other parameters, and simulated disease progression of mock PDS and NACT patients (N=50 each). Survival curves for PDS and NACT regimens within each combination of parameter values were compared based on the hazard ratio of the two groups, which was calculated by fitting a Cox proportional hazards regression model^68^ to the simulated survival data using the R *survival* package.

### Alternative Scenario 1 – Heterogeneous Chemo-resistant Cells in a Patient

We initially designed a two-component model where a single type of chemo- resistant cell exists in a patient. We then extended our mathematical framework to include a scenario in which heterogeneous chemo-resistant cells co-exist in a patient. Within this framework (Figure S3d), ovarian cancer starts from a chemo-sensitive cell with division rate *r* and death rate *d*. At each division, a chemo-sensitive cell has probability *u_1_* to convert into a Type-1 chemo-resistant cell, which divides at rate *a_1_* and dies at rate *b_1_*, and probability *u_2_* to convert into a Type-2 chemo-resistant cell, which divides at rate *a_2_* and dies at rate *b_2_*. During chemotherapy, chemo-sensitive cells divide at rate *r’* and die at rate *d’*, Type-1 chemo-resistant cells divide at rate *a_1_*’ and die at rate *b_1_*’, Type-2 chemo-resistant cells divide at rate *a_2_’* and die at rate *b_2_’*, and conversion rates are *u_1_’* and *u_2_’*, respectively. We assume that *u_1_* and *u_2_* are small enough so that conversions into Type-1 and Type-2 cells do not take place in the same division, and that the majority of cancer cells at diagnosis are chemo-sensitive.

We could then approximate the expected the numbers of Type-1 *(Y_1d_)* and Type-2 *(Y_2d_)* chemo-resistant cells at diagnosis:

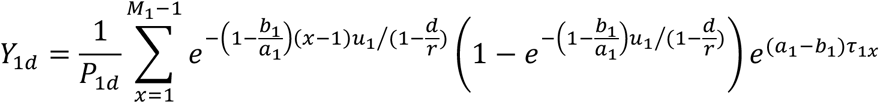

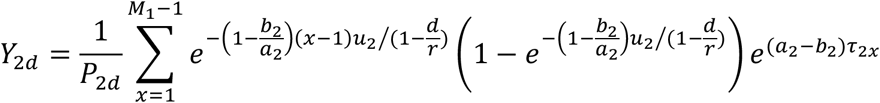

In the above equations, *τ_1x_* and *τ_2x_* represent the time from the emergence of a successful Type-1 or Type-2 chemo-resistant cell to diagnosis of disease, respectively, and approximately satisfy:

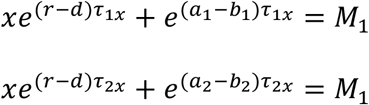

*P_1d_* and *P_2d_* represent the probabilities that Type-1 or Type-2 chemo-resistant cells exist at diagnosis, respectively, and can be expressed as:

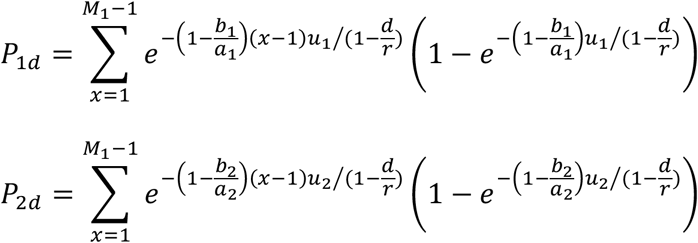

We then approximated the dynamics of chemo-sensitive (X), Type-1 chemo- resistant (*Y_1_*), and Type-2 chemo-resistant (*Y_2_*) cells as below:

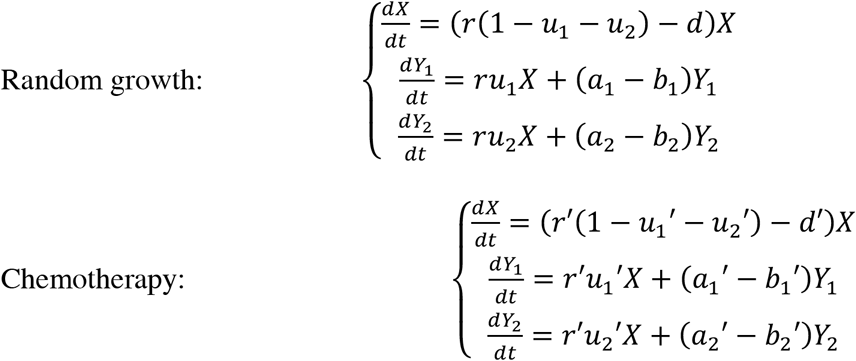

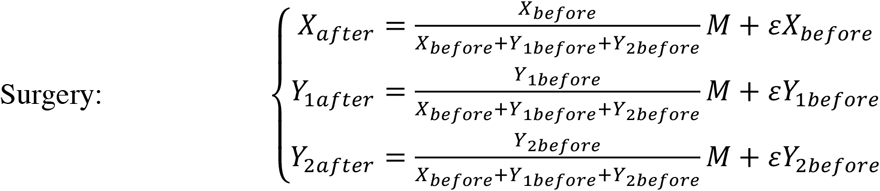

### Alternative Scenario 2 - Cancer Stem Cell Model

We also considered the alternative scenario in which HGSC originates from an intrinsically chemo-resistant cancer stem cell, which can differentiate into chemo- sensitive cancer progenitor cells^16,17,69^. Within this framework (Figure S3g), HGSC starts from a chemo-resistant stem cell with division rate *a* and death rate *b* per time unit. Each stem cell division has a probability of *u* to produce a chemo-sensitive differentiated cell, with division rate *r* and death rate *d* per time unit. During chemotherapy, chemo-resistant stem cells have division rate *a’* and death rate *b*’, and chemo-sensitive differentiated cells have division rate *r’* and death rate *d’*. Definitions of *ε, M, M_1_*, and *M_2_* are the same as in the initial model. The dynamics of chemo-resistant stem cells *(Y)* and chemo-sensitive differentiated cells can be written as:

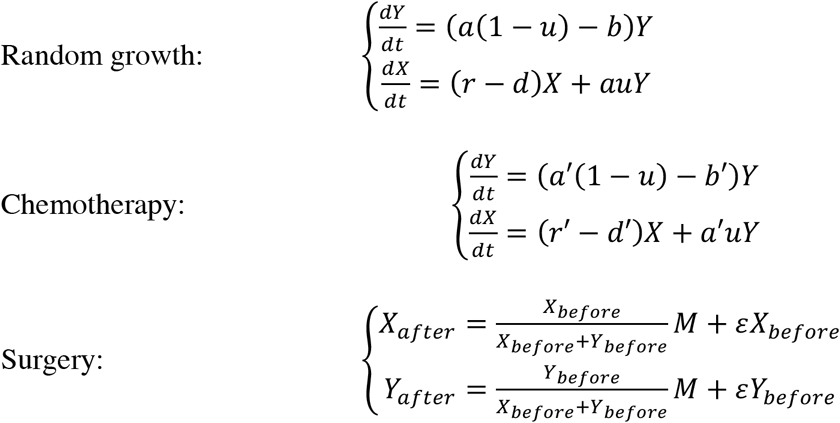

### Potential Benefit of Earlier Diagnosis of Relapsed Cancer

In order to assess the potential benefit of earlier diagnosis of relapsed cancer more stringently, we mathematically deduced and compared the expected the number of chemo-sensitive and chemo-resistant cells after early or standard diagnosis. For early diagnosis, we assumed that a patient receives chemotherapy from time 0 to *T_C_*, and the cancer randomly grows from time *T_C_* to *T_C_* + *T_R_*. For standard diagnosis, we considered that the cancer randomly grows from time 0 to *T_R_*, and chemotherapy takes place from *T_R_* to *T_R_ T_C_. T_R_* here represents the “lead time” for early diagnosis. The initial numbers of chemo-sensitive and chemo-resistant cells at the time of early diagnosis are denoted as *X_0_* and *Y_0_*, respectively.

We first modeled the effects of early diagnosis. From time 0 to *T_C_*, during chemotherapy, the dynamics of chemo-sensitive and –resistant cells can be written as:

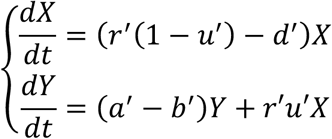

Solving these differential equations, we have:

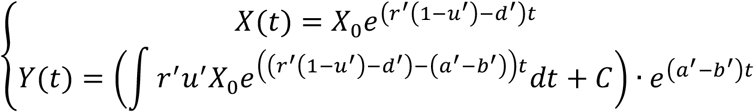

At time *T_C_*, the numbers of chemo-sensitive and –resistant cells are:

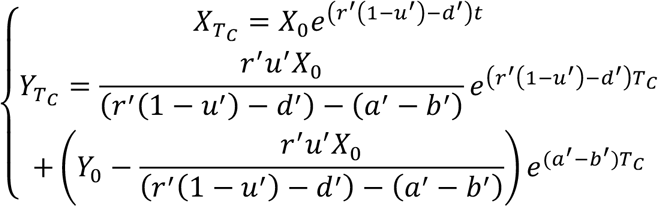

Then, from time *T_C_* to *T_C_* + *T_R_*, the dynamics of chemo-sensitive and –resistant cells can be written as:

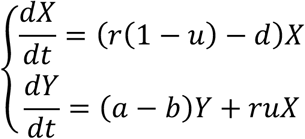

Integrating over time, we derived the numbers of chemo-sensitive and –resistant cells at time *T_C_* + *T_R_*:

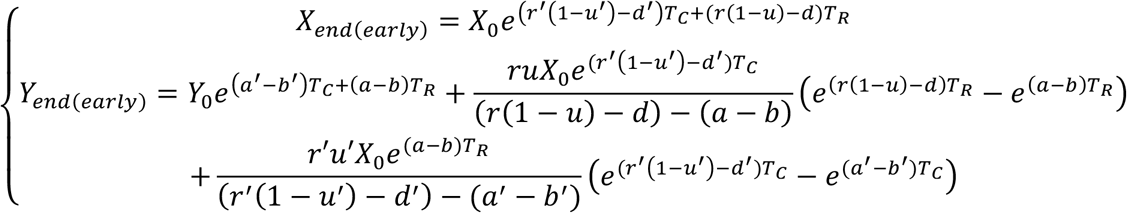

In the equation for the number of chemo-resistant cells, the term 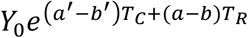 represents the number of chemo-resistant cells derived from exponential expansion of those already present at time 0, whereas the other terms represent the number of chemo-resistant cells generated from chemo-sensitive cells during the period of *T_C_* + *T_R_*.

Using similar methodology, we could deduce the numbers of chemo-sensitive and –resistant cells at *T_C_* + *T_R_* for standard diagnosis:

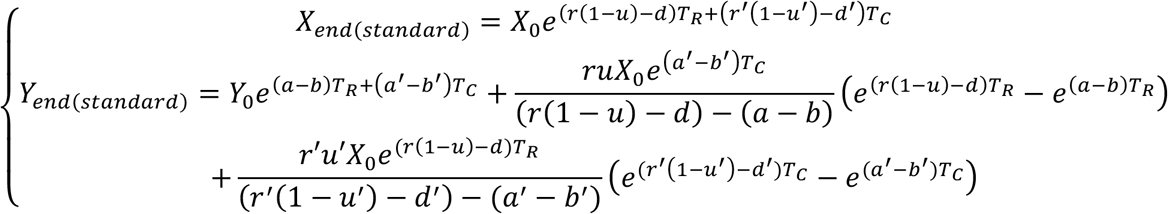

We found that *X_end_*_(_*_early_*_)_ *= X_end(standard)_*, indicating that one round of early diagnosis is unlikely to affect the number of chemo-sensitive cells. The effect on chemo-sensitive cells in Figure 6e is caused by the shorter time lapse between different lines of therapy, which is facilitated by early diagnosis.

We then calculated the difference between *Y_end_*_(_*_early_*_)_ and *X_end(standard)_:*

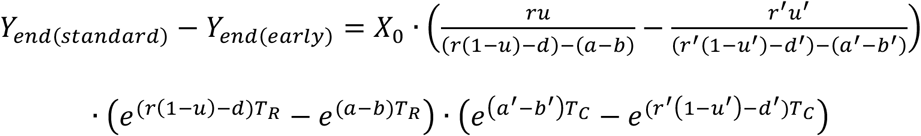

We imputed the clinically relevant values for these parameters (same as the mean values shown in Figure 2), and interrogated the influence of *T_R_* on the relative difference in chemo-resistant cell numbers between early and late diagnosis:

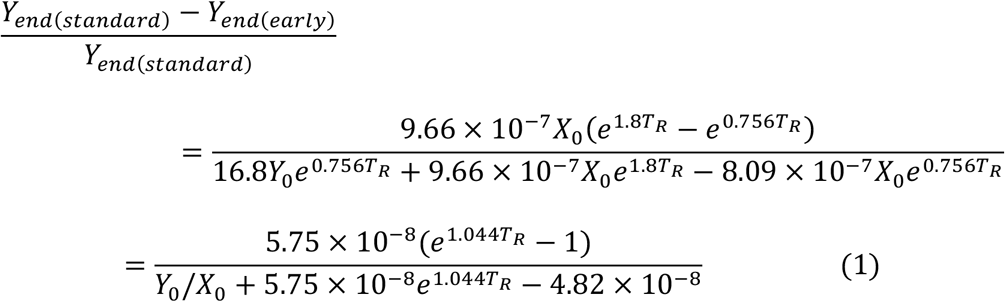

Equation (1) demonstrates that the relative difference in chemo-resistant cells is associated with two factors: (1) the initial ratio of chemo-resistant/-sensitive cells at early diagnosis (++*Y_0_*/ *X_0_*), and (2) the length of lead time for early diagnosis (TR). If we impute *Y_0_*/*X_0_*=0.01, meaning ~1% cancer cells at early diagnosis of relapsed cancer are chemo-resistant, then *T_R_* needs to be longer than 11.6 months in order for the ratio in equation (1) to be less than 0.5, corresponding to an advantage of ~1 month in overall survival for early diagnosis. If 0.1% cancer cells at early diagnosis of relapsed cancer are chemo-resistant, then *T_R_* needs to be longer than 9.4 months for an advantage of ~1 month in overall survival.

### Estimation of Conversion Rate *u* and Unresectable Proportion *ε*

For each combination of candidate values for *u* and *ε*, we calculated the deviation of model prediction from clinical observations as:

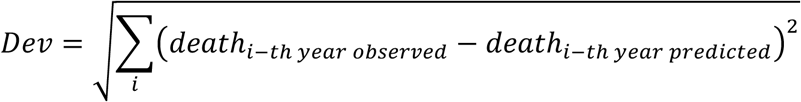

The combination that led to lowest deviation was used for model testing and further predictions.

### Statistical Analyses

Comparisons of overall survival between 2 groups were performed by log-rank test. Comparisons of predicted survival between >3 groups were performed by log-rank test for trend when the order of groups is logical. Comparisons of distribution of overall survival between model predictions and clinical observations were performed by Chi-square test.

## Data Availability

All data were acquired from published clinical trials or retrospective studies.

## Author Contributions

S.G. and B.G.N. designed the study; S.G. constructed the computational model and performed the analyses; S.L., L.H.B., I.V., P.C., M.B., B.R., and A.O. provided clinical expertise and helpful discussions; A.S. provided computational and statistical expertise and helpful discussions; L.H.B. and I.V. extracted raw clinical data from UHN dataset; *d*.T. and W.P. extracted raw clinical data from NCIC dataset; and B.G.N. supervised the research. S.G. and B.G.N. wrote the manuscript.

## Acknowledgements

The authors thank Dr. Hiroshi Haeno from Kyushu University and Drs. Siv Sivaloganthan and Mohammad Kohandel from Waterloo University for helpful discussions. This work was supported by grants from the Terry Fox Foundation (TFPPG 020003), National Cancer Institute (R37 49152), and Department of Defense (60167542 104703 A) (to B.G.N.), and a DCA Award from University of Toronto (to S.G.). B.G.N. was a Canadian Research Chair, Tier 1, and work in his laboratory was supported in part by a grant from the Ontario Ministry of Health and Long Term Care and the Princess Margaret Cancer Foundation.

---

**Figure S1.**
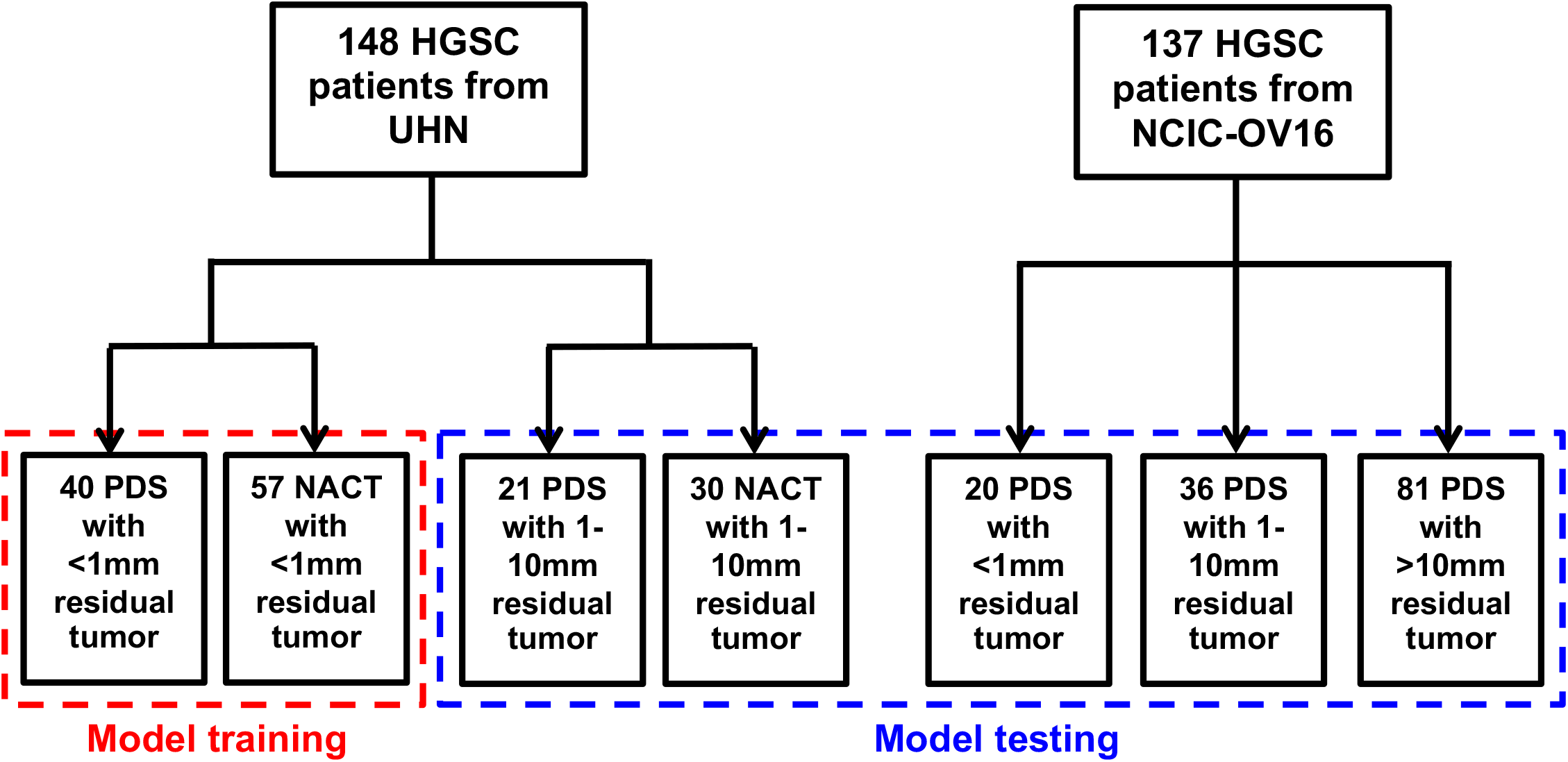
Data Organization for Model Training and Testing.

**Figure S2.**
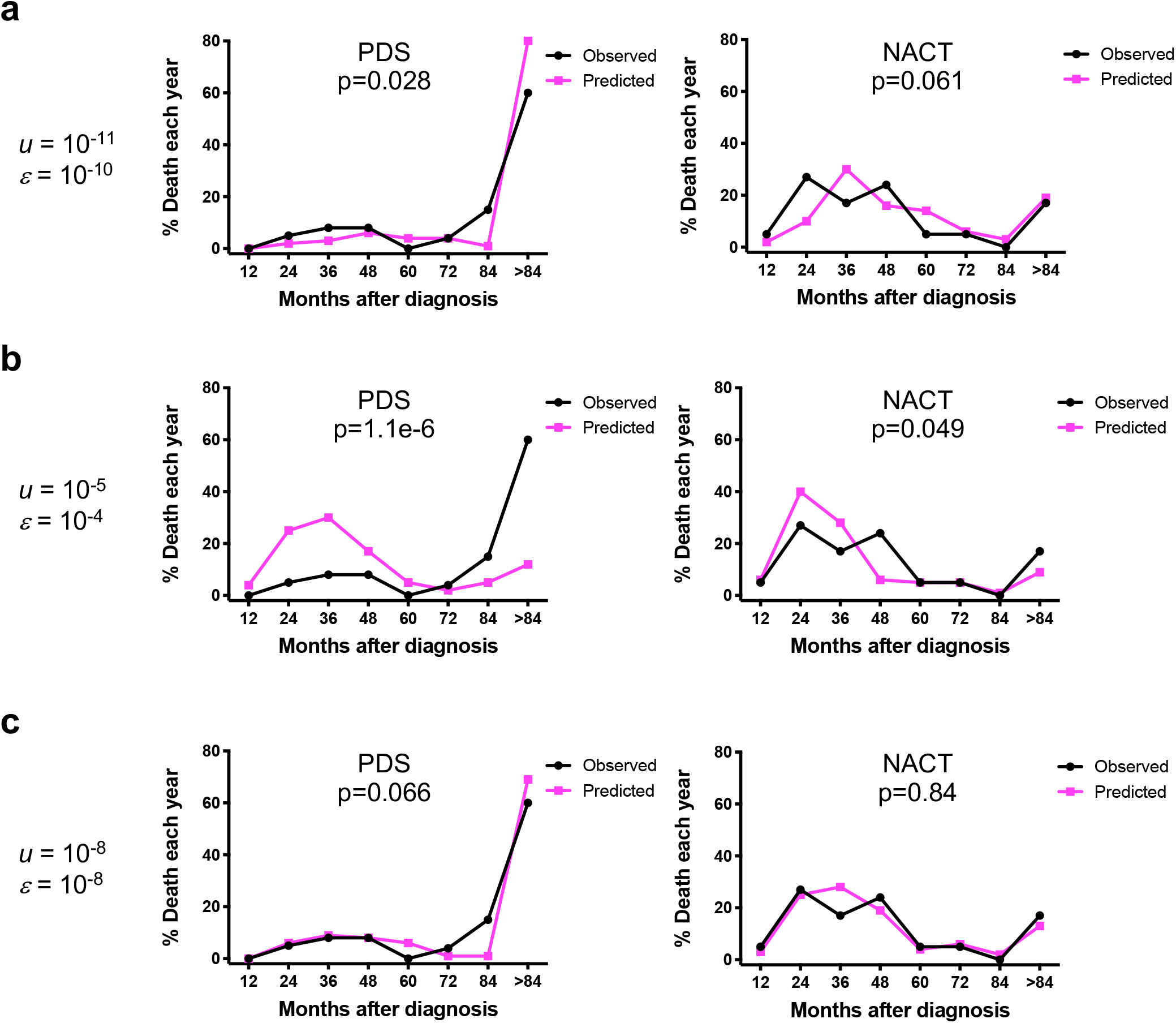
Evaluation of alternative *u* and *ε* values with training set data.

**Figure S3.**
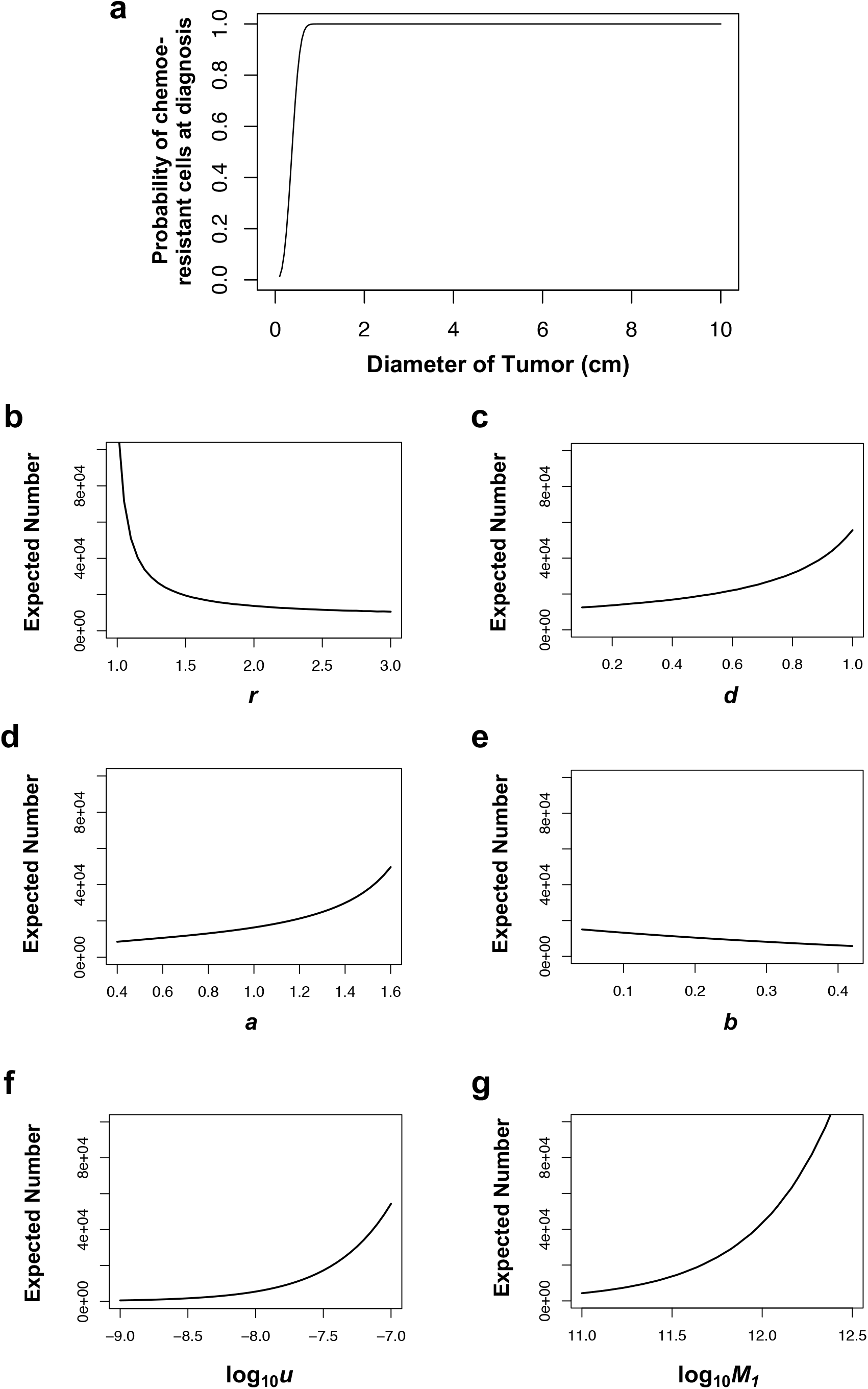
Dependence of Number of Resistant Cells at Diagnosis on Parameter Values.

**Figure S4.**
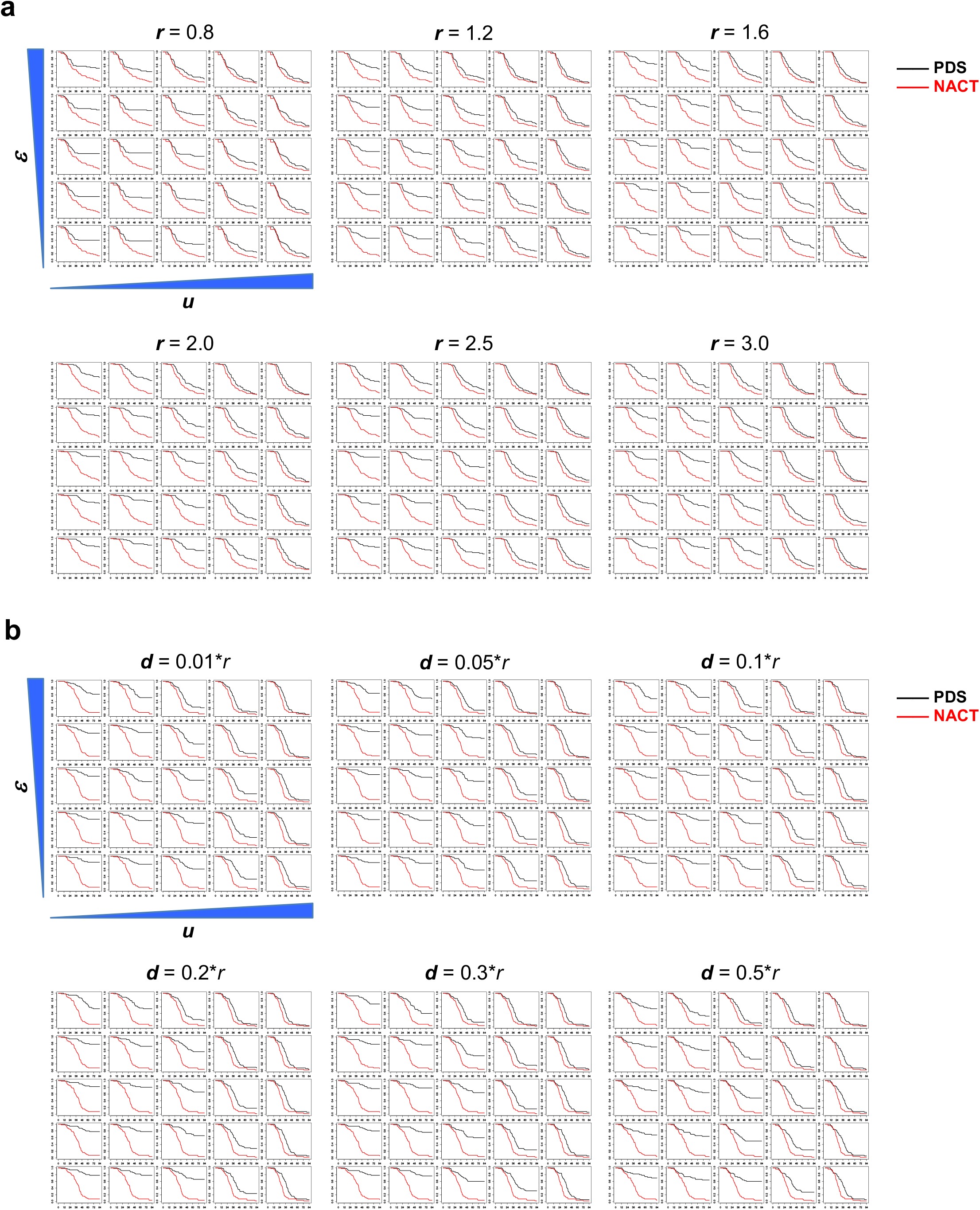
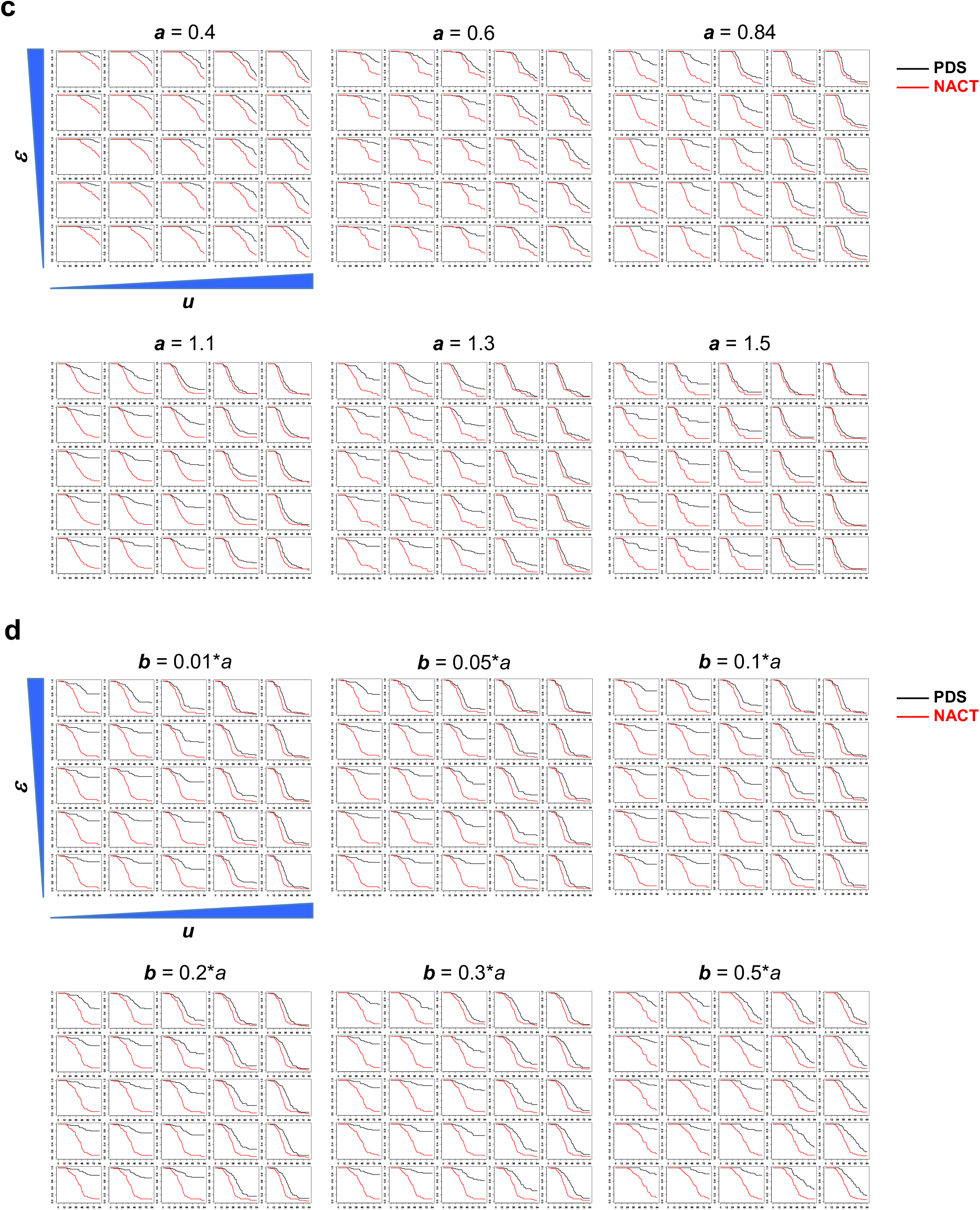
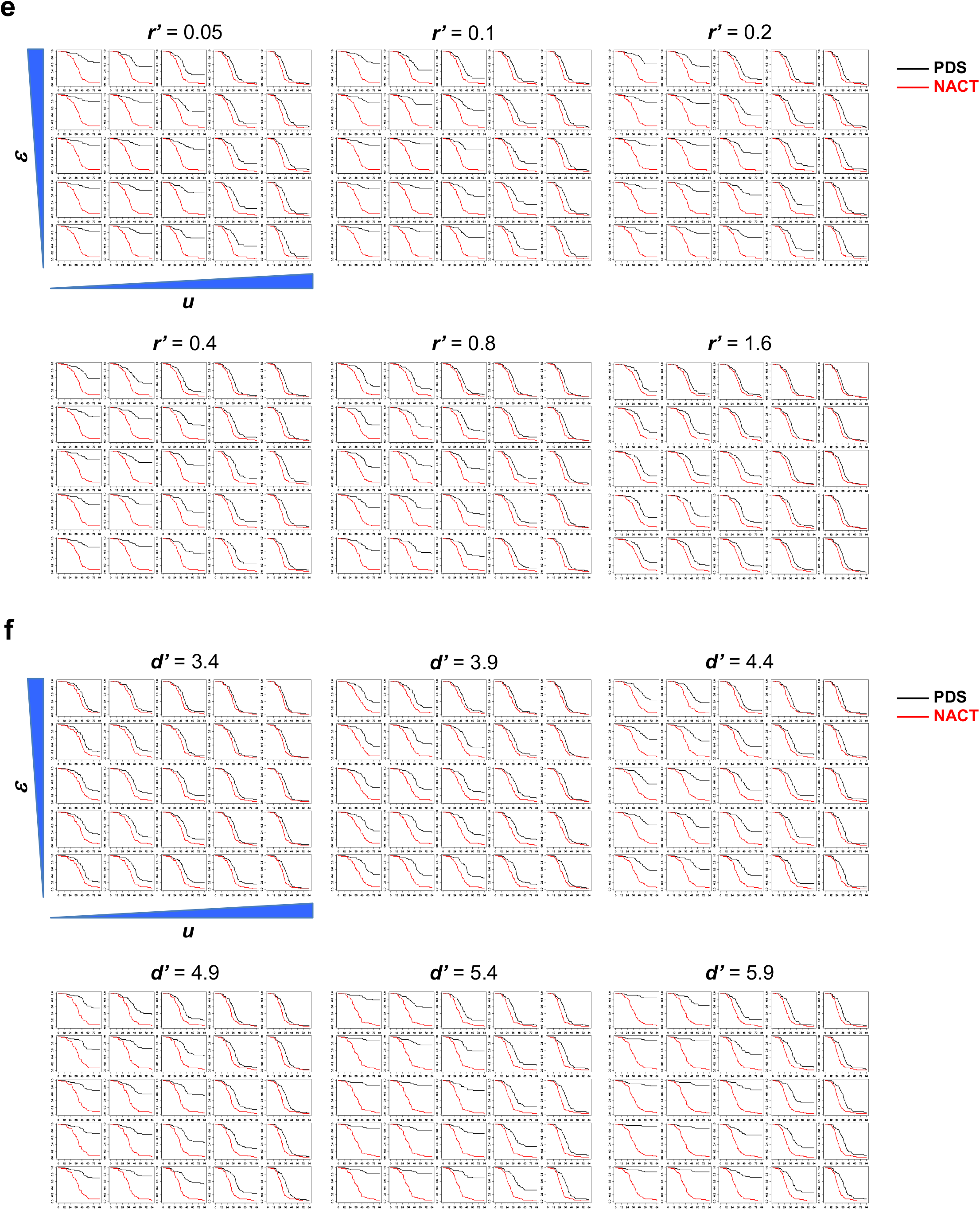
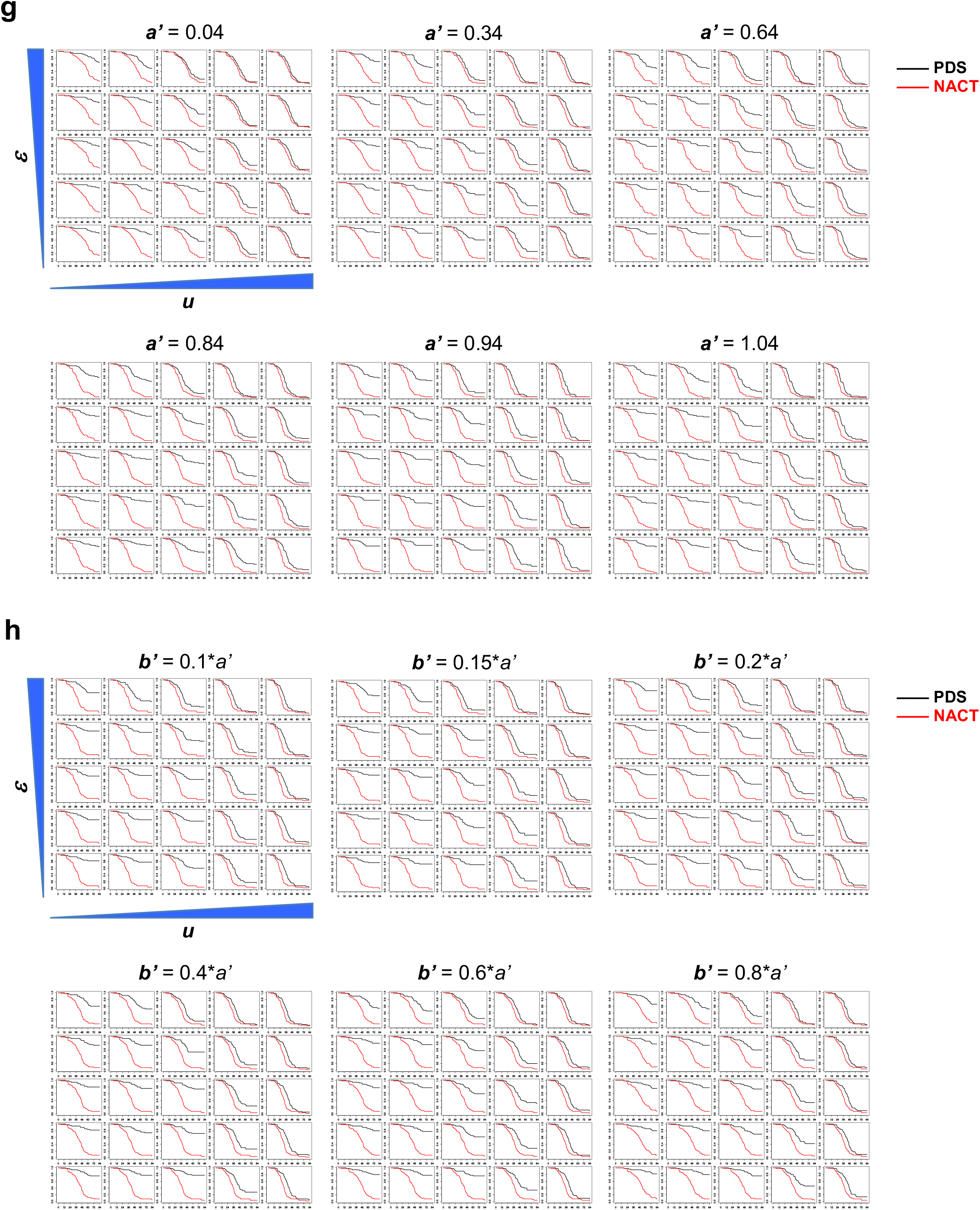
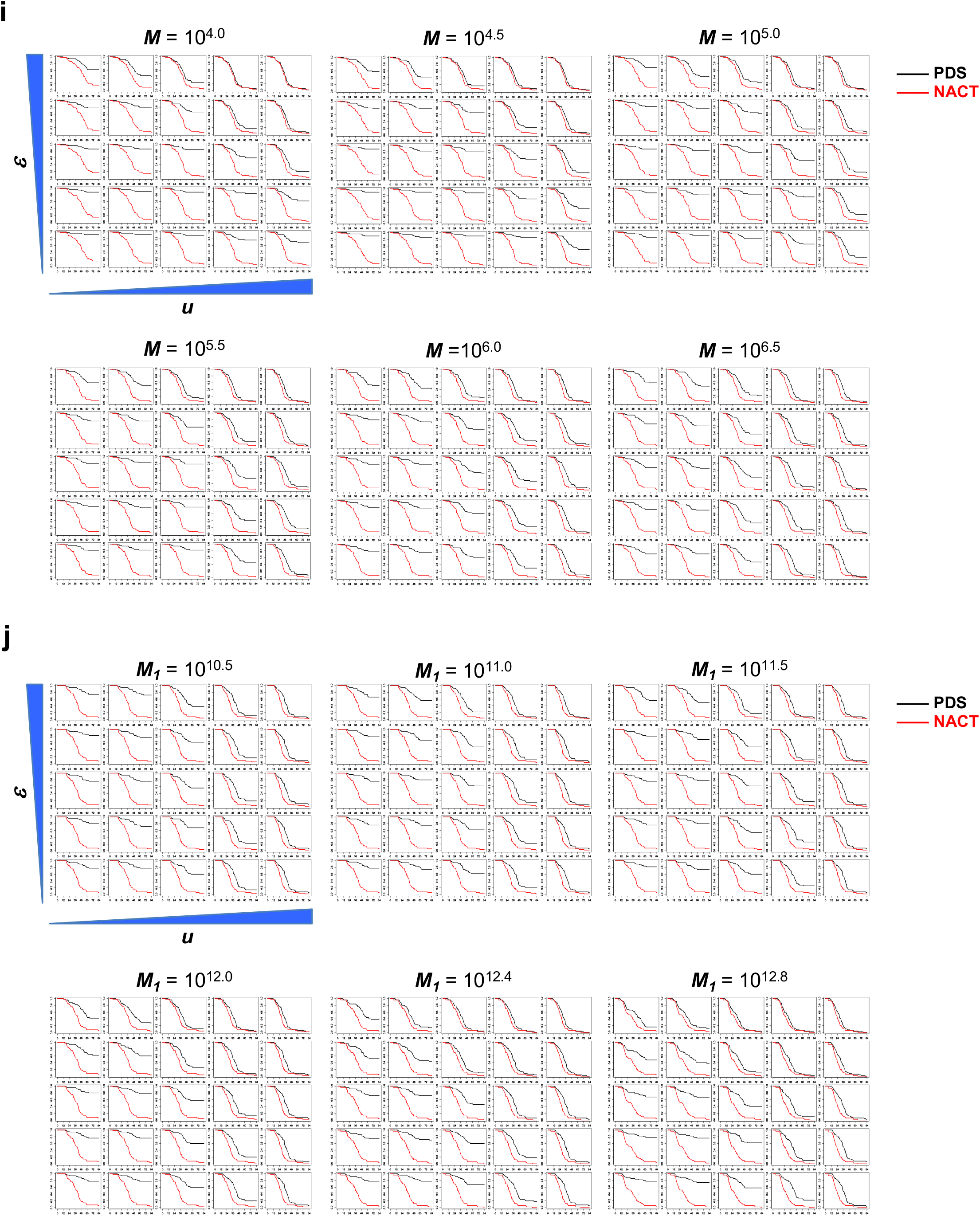
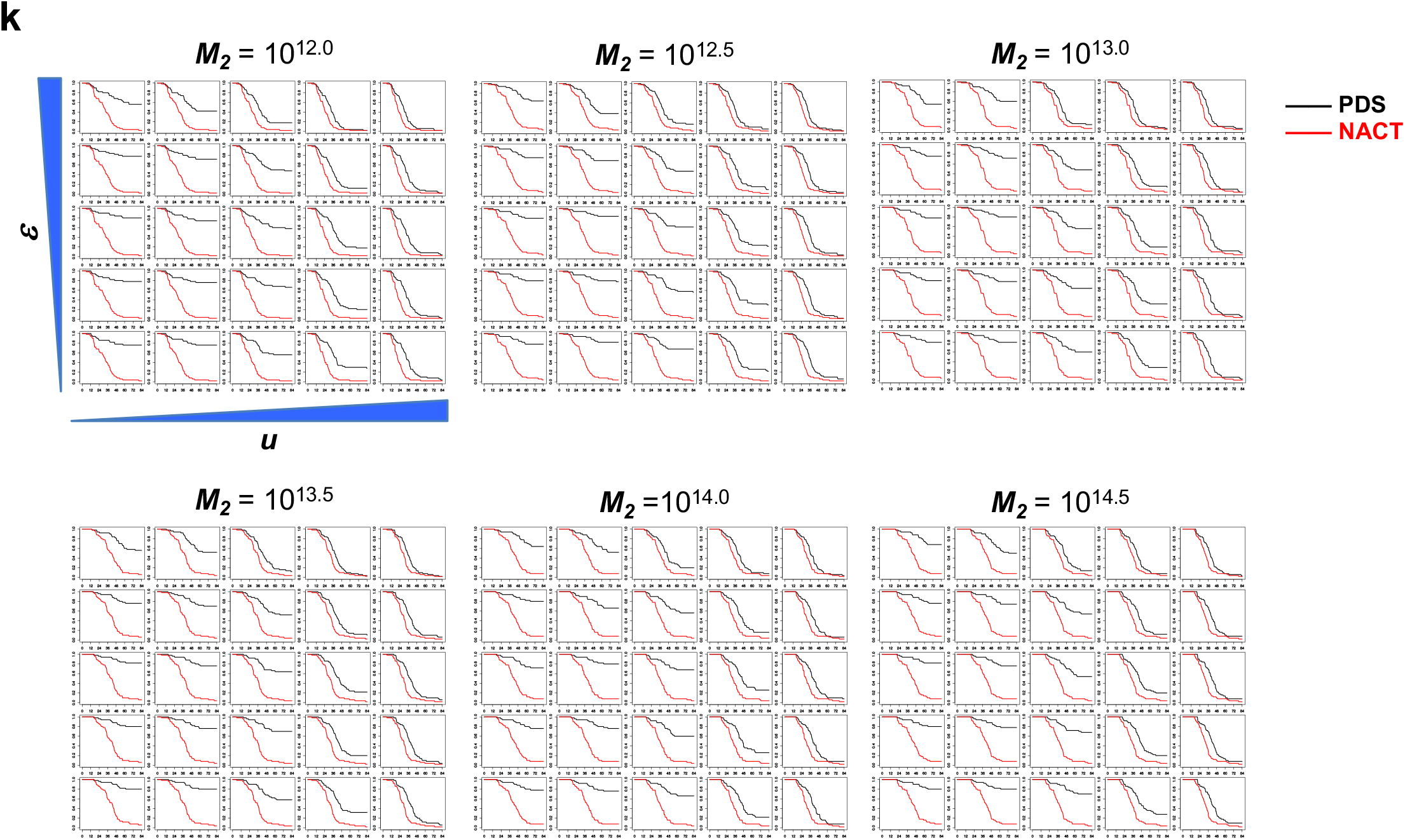
Calculated Effects of Varying Parameter Values on Survival Following PDS or NACT.

**Figure S5.**
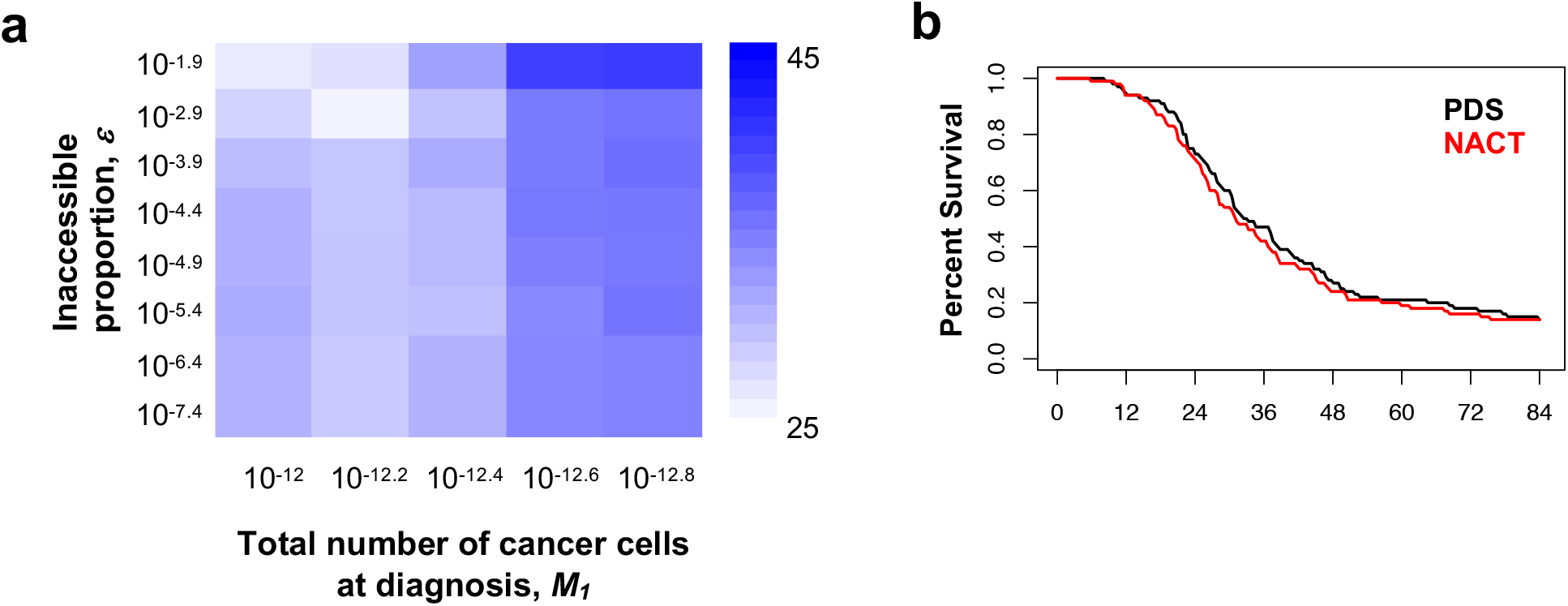
Comparison of PDS and NACT using parameters trained from the CHORUS trial.

**Figure S6.**
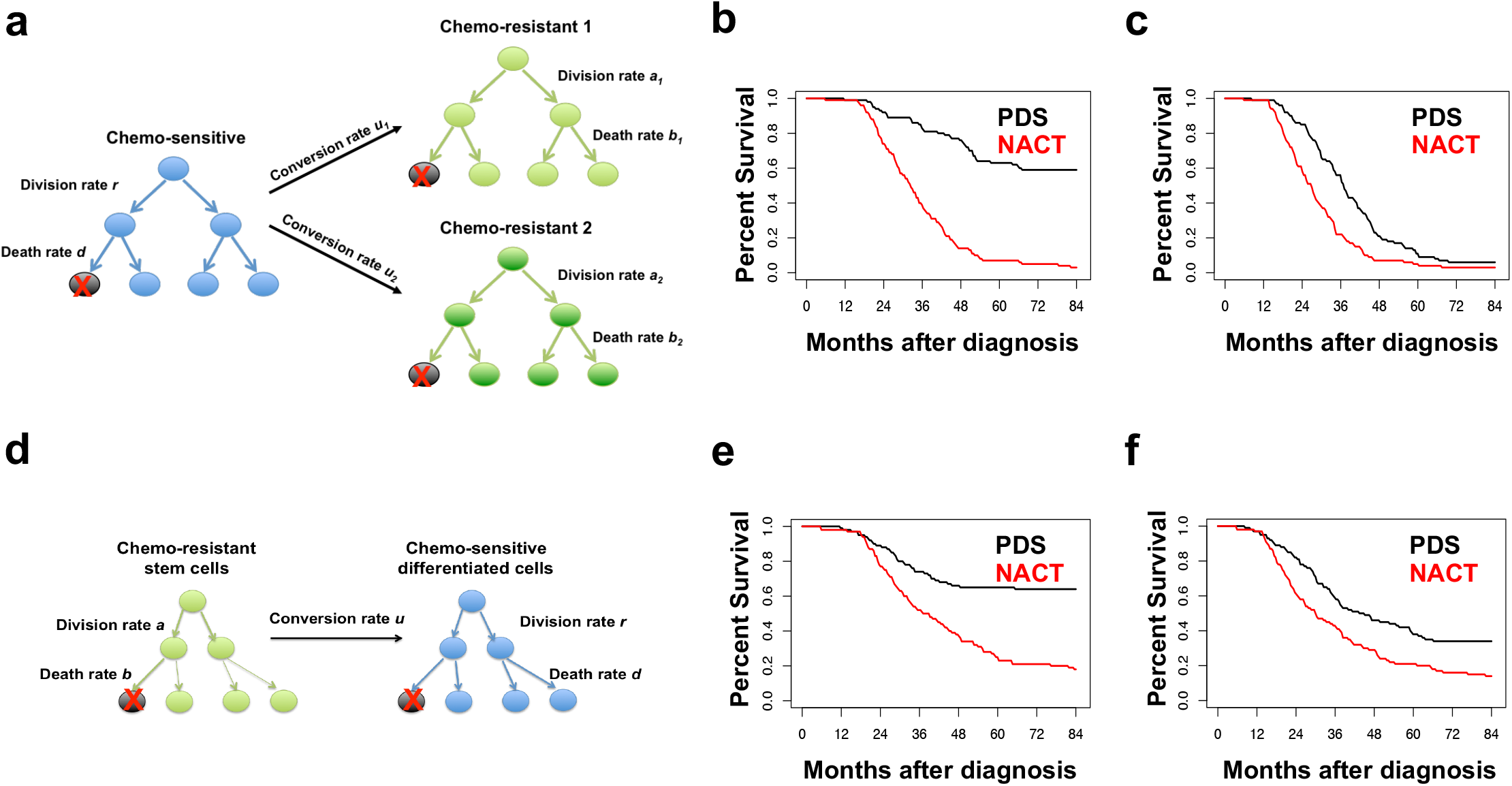
Calculated Effects of Varying Model Assumptions on Survival Following PDS or NACT.

**Figure S7.**
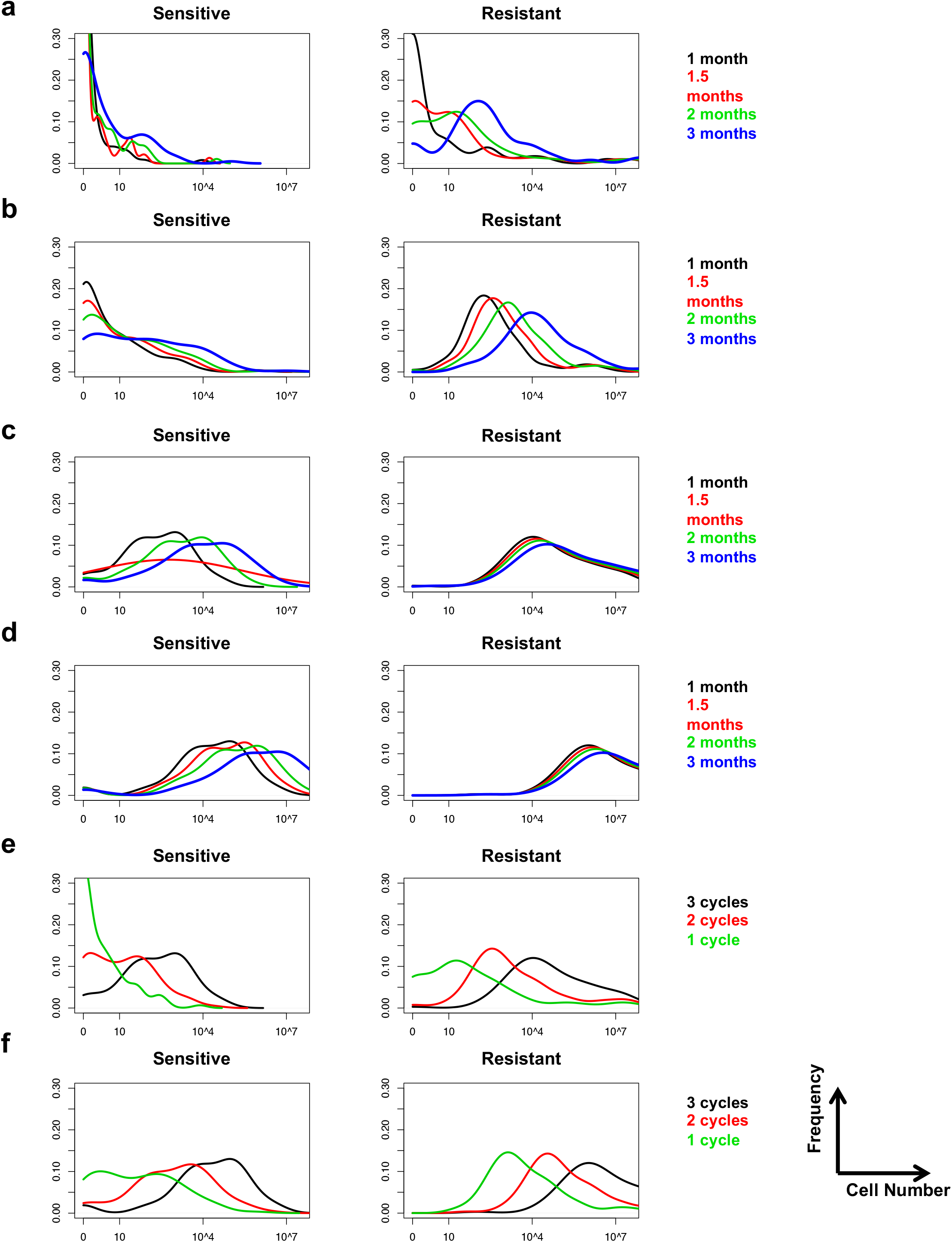
Residual Tumor Cells after Different Treatment Strategies.

**Figure S8.**
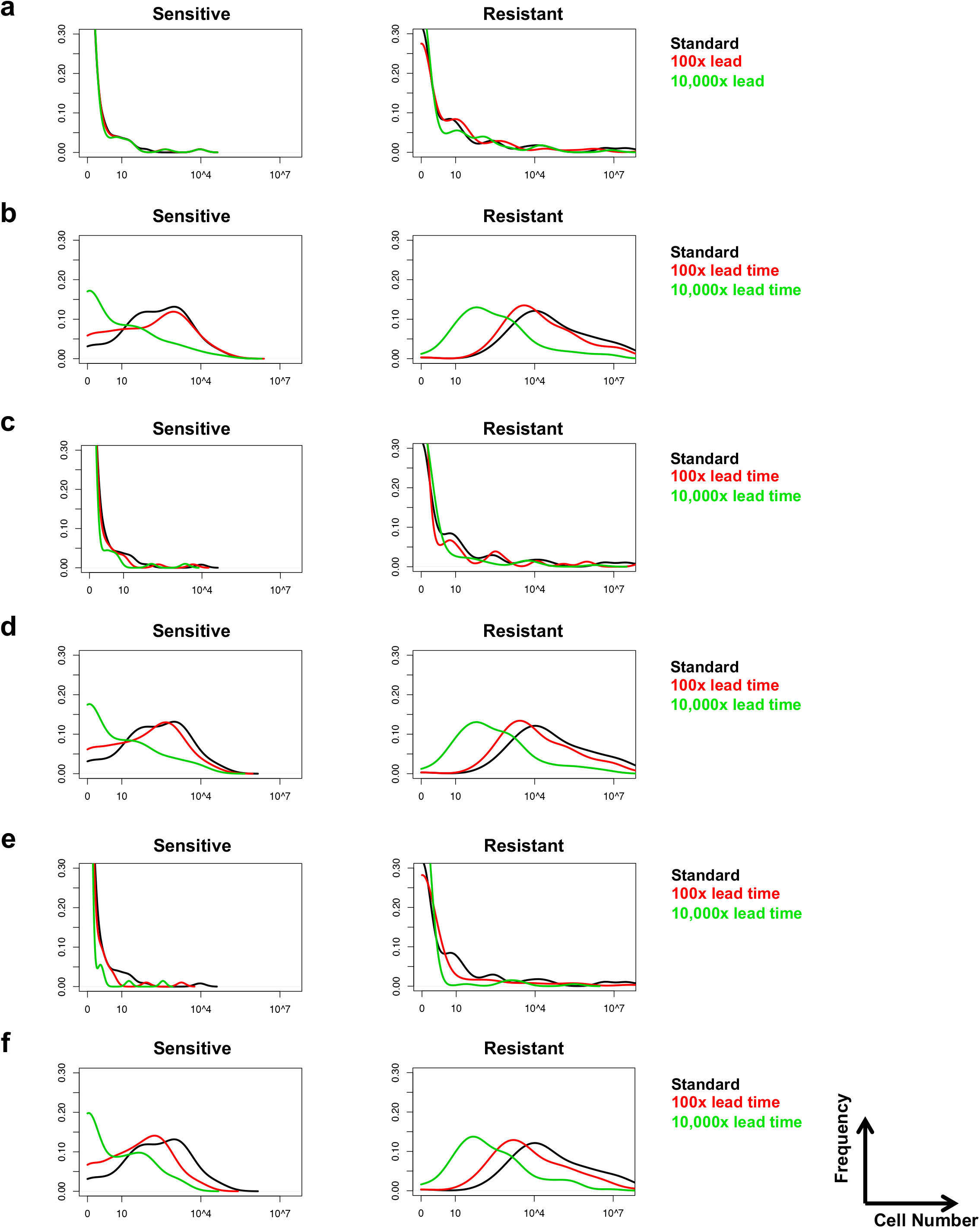
Tumor Cells Remaining after First-Line Therapy with Earlier Diagnosis.

